# Systems biological assessment of the temporal dynamics of immunity to a viral infection in the first weeks and months of life

**DOI:** 10.1101/2023.01.28.23285133

**Authors:** Florian Wimmers, Allison R. Burrell, Yupeng Feng, Hong Zheng, Prabhu S. Arunachalam, Mengyun Hu, Sara Spranger, Lindsay Nyhoff, Devyani Joshi, Meera Trisal, Mayanka Awasthi, Lorenza Bellusci, Usama Ashraf, Sangeeta Kowli, Katherine C. Konvinse, Emily Yang, Michael Blanco, Kathryn Pellegrini, Gregory Tharp, Thomas Hagan, R. Sharon Chinthrajah, Alba Grifoni, Alessandro Sette, Kari C. Nadeau, David B. Haslam, Steven E. Bosinger, Jens Wrammert, Holden T. Maecker, Paul J. Utz, Taia T. Wang, Surender Khurana, Purvesh Khatri, Mary A. Staat, Bali Pulendran

**Affiliations:** Institute for Immunity, Transplantation and Infection, Stanford University, Stanford, CA, USA; Department of Molecular Medicine, Interfaculty Institute for Biochemistry, University of Tuebingen, Tuebingen, Germany; DFG Cluster of Excellence 2180 ‘Image-guided and Functional Instructed Tumor Therapy’ (iFIT), University of Tuebingen, Tuebingen, Germany; German Consortium for Translational Cancer Research (DKTK), German Cancer Research Center (DKFZ), Heidelberg, Germany; Department of Infectious Diseases, Cincinnati Children’s Hospital Medical Center, Cincinnati, OH, USA; Department of Environmental and Public Health Sciences, Division of Epidemiology, University of Cincinnati College of Medicine, Cincinnati, OH, USA; Center for Biomedical Informatics Research, Department of Medicine, Stanford University, Stanford, CA, 94305, USA; Department of Pediatrics, Division of Infectious Disease, Emory University School of Medicine; Division of Viral Products, Center for Biologics Evaluation and Research, Food and Drug Administration, Silver Spring, Maryland, 20993, USA; Department of Medicine, Division of Infectious Diseases, Stanford University, Stanford, CA 94305, USA; Department of Microbiology and Immunology, Stanford University School of Medicine, Stanford University, Stanford, CA, USA; Department of Pediatrics, Stanford University School of Medicine, Stanford University, Stanford, CA, USA; Department of Medicine, Division of Immunology and Rheumatology, Stanford University School of Medicine, Stanford, CA, USA; Stanford Genomics Service Center, Department of Genetics, Stanford University School of Medicine, Stanford, CA, USA; Yerkes National Primate Research Center, Atlanta, GA, USA; Department of Pediatrics, University of Cincinnati College of Medicine, Cincinnati, OH, USA; Department of Medicine, Sean N. Parker Center for Allergy and Asthma Research, Stanford, CA 94305, USA; Center for Infectious Disease and Vaccine Research, La Jolla Institute for Immunology (LJI), La Jolla, CA 92037, USA; Department of Medicine, Division of Infectious Diseases and Global Public Health, University of California, San Diego, La Jolla, CA 92037, USA; Department of Pathology, Emory University School of Medicine, Atlanta, GA, USA; Department of Pathology, Stanford University School of Medicine, Stanford University, Stanford, CA, USA

## Abstract

The dynamics of innate and adaptive immunity to infection in infants remain obscure. Here, we used a multi-omics approach to perform a longitudinal analysis of immunity to SARS-CoV-2 infection in infants and young children in the first weeks and months of life by analyzing blood samples collected before, during, and after infection with Omicron and Non-Omicron variants. Infection stimulated robust antibody titers that, unlike in adults, were stably maintained for >300 days. Antigen-specific memory B cell (MCB) responses were durable for 150 days but waned thereafter. Somatic hypermutation of V-genes in MCB accumulated progressively over 9 months. The innate response was characterized by upregulation of activation markers on blood innate cells, and a plasma cytokine profile distinct from that seen in adults, with no inflammatory cytokines, but an early and transient accumulation of chemokines (CXCL10, IL8, IL-18R1, CSF-1, CX3CL1), and type I IFN. The latter was strongly correlated with viral load, and expression of interferon-stimulated genes (ISGs) in myeloid cells measured by single-cell transcriptomics. Consistent with this, single-cell ATAC-seq revealed enhanced accessibility of chromatic loci targeted by interferon regulatory factors (IRFs) and reduced accessibility of AP-1 targeted loci, as well as traces of epigenetic imprinting in monocytes, during convalescence. Together, these data provide the first snapshot of immunity to infection during the initial weeks and months of life.

## Introduction

Infants and young children are born with an immune system that differs in composition and functionality from adults ^1–3^ and undergoes profound maturation during the initial weeks and months of life ^1, 3^. While previous studies have described this maturation process in healthy infants ^1^, a detailed system-wide, longitudinal analysis of the immune response to an infection in infants has yet to be undertaken. Here, we address this knowledge gap by assessing immunity to SARS-CoV-2 early after birth. In contrast to adults, infants and children develop mild symptoms after infection ^4^, although severe cases and deaths have been observed ^5^. While previous publications primarily described immune responses to COVID-19 in older children (median age five years) with a relatively mature immune system ^6–9^, little is known about how the immature immune system responds to SARS-CoV-2 infection during the first weeks and months of life. Several key questions arise in this context: 1) Given the nascency of the adaptive immune system in this age group ^2, 3^, to what extent do infants and young children develop durable antibody responses and T and B cell memory to the SARS-CoV-2 virus? 2) In light of the mild course of pediatric COVID-19, what are the hallmarks of innate immune activation compared to that observed in adults? 3) Studies in older children and adults reported autoantibodies and lasting epigenomic changes after COVID-19 ^10–12^. How does SARS-CoV-2 infection impact the maturing infant immune system in the long term? To answer these questions, we used a multi-omics approach and profiled immunity to SARS-CoV-2 infection in a longitudinal cohort of infants and young children during the first weeks and months of life.

## Results

### Study cohort

We obtained pediatric COVID-19-infected, and healthy control samples from infants and young children enrolled in the IMPRINT cohort at the Cincinnati Children’s Hospital Medical Center. All infants and young children were tested weekly for SARS-CoV-2 and healthy controls tested negative from birth to sampling. Overall, we analyzed 125 samples from 54 infected and 27 healthy infants and young children (**Figure 1a**). Our cohort contains samples from infants and young children infected with different SARS-CoV-2 variants: 32 infants and young children were infected with pre-Omicron variants, and 22 were infected with Omicron variants (**Figure 1a, DataS1**). Samples in the pre-Omicron cohort were collected longitudinally, with paired samples from before, during, and after infection (**Figure 1a**). The age at infection was 1 to 47 months (median age 9 months), and 56% of pediatric patients were male (**DataS1**). In addition, we obtained 62 samples from 48 adult COVID-19 patients and ten healthy controls from the Hope Clinic at Emory University in Atlanta and the Stanford University Medical Center (**DataS1**). The median age in the adult cohort was 59 years; 48% of adult patients were male. Details on patient demographics, disease severity, and assay distribution can be found in Supplementary Materials (**DataS1; Supp Figure 1a**).

**Figure 1.**
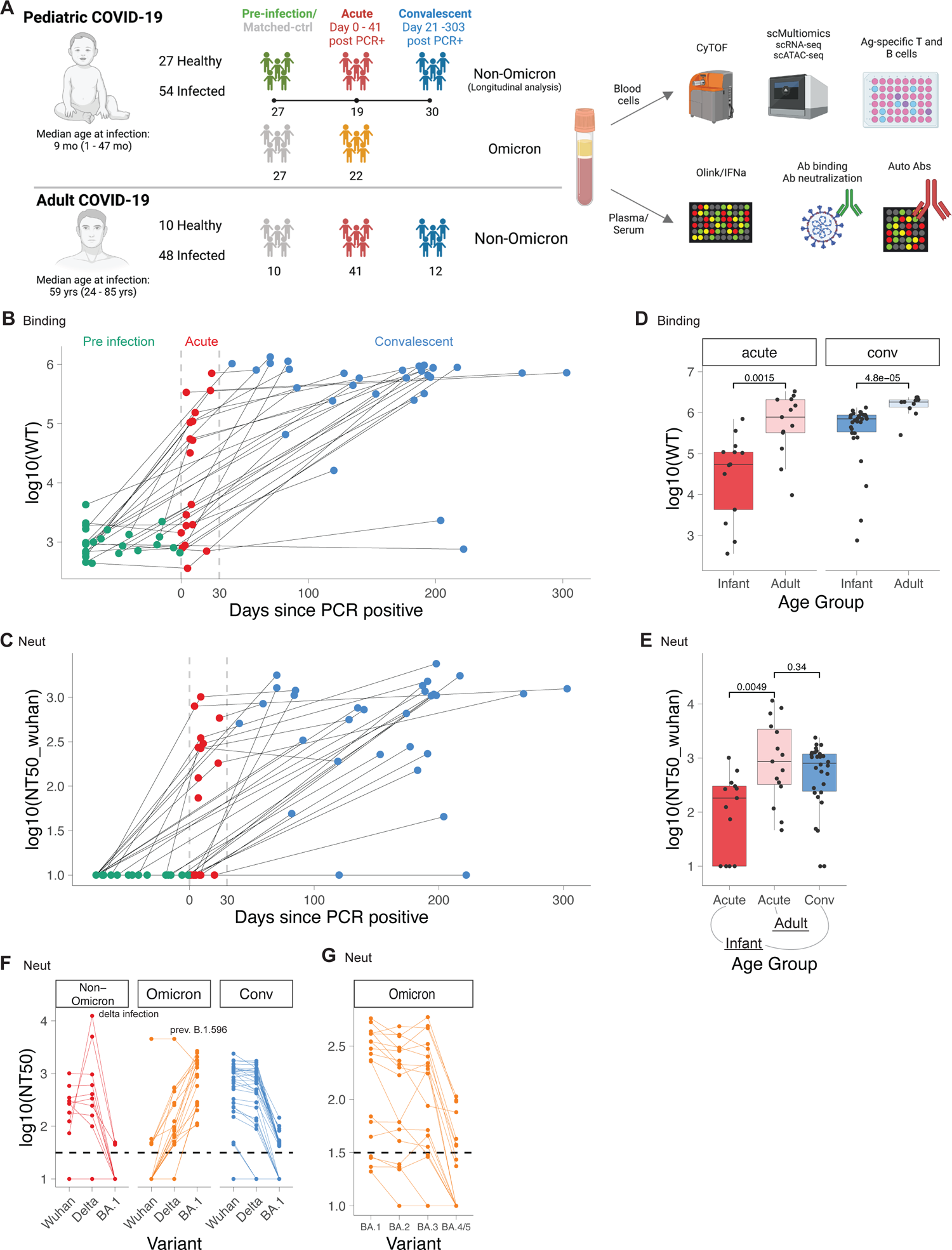
Durable antibody response in infants and young children with COVID-19. A) Study layout. B-G) Antibody binding and neutralization titers of infants and adults with COVID-19. B) Antibody binding titers to WT strain in longitudinal samples from infants taken before (Pre, n = 27), during (Acute, n = 19), and after (Conv, n = 30) infection. Dotted lines indicate days 0 and 30 post PCR+. C) Line graph showing neutralizing antibody titers to Wuhan strain. D) Binding titers to WT strain in adults (acute, n = 13; Conv = 10) and infants (acute n = 13, conv = 30). E) Neutralizing titers to Wuhan strain in adults (acute n = 15) and infants (acute n = 13, conv n = 30). F, G) Neutralizing titers against different variants in infants (Non-Omicron n = 13, Omicron n = 18, Conv n = 30). D-G) Shown are only samples > 5 days post infection. Statistical comparisons with Wilcoxon rank sum test.

### Robust and persistent antibody response

Whilst recent studies have documented the kinetics of the antibody response to SARS-CoV-2 infection in adults, there is currently no such information in the infant population. The availability of samples obtained before, during, and following SARS-CoV-2 infection permitted us to perform a longitudinal analysis of the magnitude and kinetics of the antibody response during the first weeks and months of life. We thus measured binding and neutralizing antibody titers against a range of SARS-CoV-2 variants, including the Omicron variants BA.1, BA.2, BA.3, and BA.4/5 using a multiplexed electrochemiluminescence assay and pseudovirus neutralization assays. For the antibody analysis, we analyzed a subset of 128 samples from 55 infants and young children and 29 adults with polymerase chain reaction (PCR)-confirmed SARS-CoV-2 infection (CoV-2+). While pre-infection binding and neutralization titers were low or nonexistent in infants and young children, respectively, anti-Spike antibodies emerged about 4-5 days after first testing CoV-2+ (**Figure 1b,c, Supp Figure 1b,c)**, in line with previous studies in adults ^13^. Strikingly, during acute infection, there was a wide variation in the magnitude of the neutralizing and binding antibody titers, with some infants and young children having undetectable responses (**Figure 1b,c, Supp Figure 1b,c**). During the convalescent phase, 28 out of 30 infants and young children developed robust antibody responses (**Figure 1b,c**), which were durably maintained for the entire observation period of more than 300 days (**Figure 1b,c)**. This contrasts with previous studies in adults that demonstrated a marked decay in the antibody response during the weeks and months following infection ^14–16^. However, the magnitude of the antibody binding titers in infants and young children was significantly lower than in adults during both acute infection and convalescence (**Figure 1d,e**).

We next assessed the breadth of the antibody response against SARS-CoV-2 variants. Infection with non-Omicron variants induced binding and neutralization antibody responses against all measured pre-Omicron variants but bound and neutralized Omicron variants poorly **(Figure 1f, Supp Figure 1d,e**). In contrast, infants and young children infected with the Omicron variant developed robust antibody binding and neutralizing titers against Omicron variants (**Supp Figure 1d,e, Figure 1f**). Notably, within the group of Omicron-infected infants and young children, subjects infected with BA.1 showed substantially reduced neutralizing titers for BA.4/5 (**Figure 1g**). Together, these data demonstrate the induction of robust and durable antibody responses against SARS-CoV-2 during the first year of life but also highlight the limited cross-neutralization ability of induced antibodies.

### Autoantibody response

Previous studies reported autoantibodies against type I IFNs as a driver of severe COVID-19 infection in adults ^11^ and autoantibodies against multiple self-antigens as a critical feature of Multisystem Inflammatory Syndrome in Children (MIS-C) ^6, 10, 17, 18^. Whether autoantibodies against cytokines and self-antigens are also a key feature of COVID-19 infection during the first weeks and months of life is unknown. To test this, we analyzed plasma samples from 77 infants and young children and 25 adults using two custom bead-based protein arrays to measure IgG antibodies found in connective tissue diseases and anti-cytokine antibodies (**DataS2**). We detected few autoreactive antibodies in pre-infection samples, while acute infection, convalescent, and matched-control samples showed autoantibodies against an extensive range of targets (**Supp Figure 1f, Supp Figure 2**). Type I IFN antibodies were sporadically detected in a small subset of infants and young children at all time points, but no infection-induced increase was evident. In contrast, a greater proportion of the infected adults contained type I IFN autoantibodies relative to healthy adults (**Supp Figure 2**). Interestingly, in many infants and young children, we observed an increase in the concentration of autoantibodies against Sm proteins B/B’ (Smith), which are the structural components of splicing complexes and are associated with systemic lupus erythematosus (SLE) (**Supp Figure 2**). During acute infection, a few infants and young children had a transient increase in specific autoantibodies associated with myositis and autoimmune overlap syndromes (MI-2, Jo1) and scleroderma (CENP A) as well as anti-cytokine antibodies (IL-17A, IL-21, CNTF, IL-1beta). Intriguingly, other autoantibodies associated with SLE (PCNA), myositis (SRP54), and scleroderma (Fibrillarin), as well as anti-cytokine antibodies (IL-33, ACE2, VEGFB, IL-12p40), were persistent or increased in convalescent samples (**Supp Figure 1f**).

**Figure 2.**
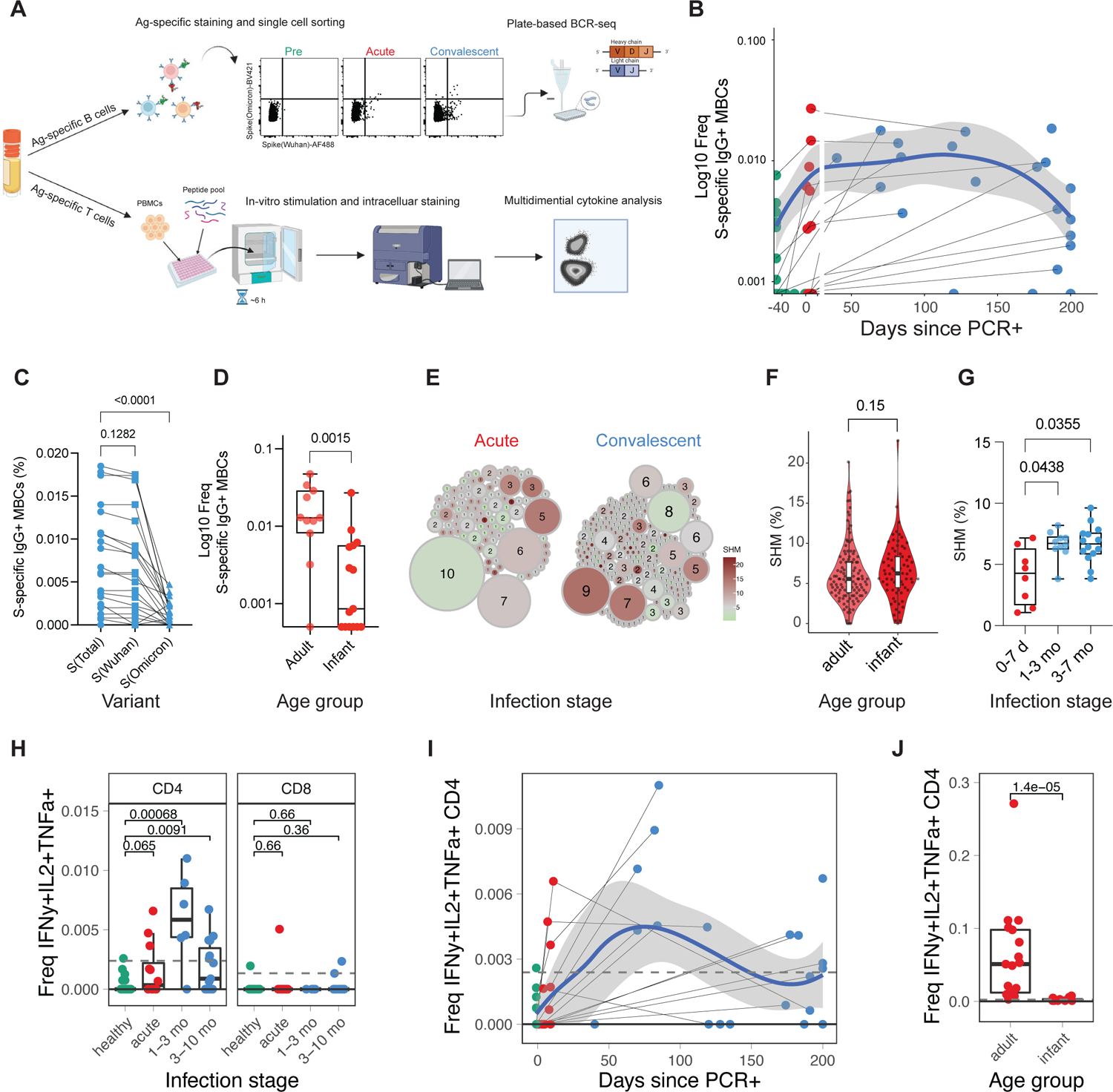
Transient memory B and T cell response to COVID-19 infection in infants and young children. A) Diagram depicting the experiment (n infants and young children: pre=12, acute=12, conv=21; n adults: acute=17). B-G) Spike-specific B cells were sorted using FACS, and the BCR sequence of each clone was determined. B) Frequency of SARS-CoV-2 spike-specific IgG+ memory B cells as a proportion of CD20+ B cells. Samples of the same donor a connected by a gray line. Blue line indicates average values; shaded areas indicate 5th to 95th percentiles. C,D) Frequency of SARS-CoV-2 specific IgG+ memory B cells in C) infants and young children at convalescent phase and D) at acute phase in adults and infants and young children. E) Clonality analysis of sorted SARS-CoV-2 spike-specific IgG+ memory B cells in infants and young children (n = 220). Each clone is represented as a circle. Circle size indicates the number of IGHV sequences in each clone; color represents the mean IGHV somatic hypermutation rate. F) Somatic hypermutation rates of the IGHV genes in single sorted SARS-CoV-2 spike specific IgG+ memory B cells at acute phase. G) Mean somatic hypermutation rate of all cloned IGHV genes in indicated infant samples. H-J) T cells were stimulated with overlapping peptides against WT and Omicron variants. Cytokine production was determined via flow cytometry. H) Box plot showing the fraction of multifunctional T cells (IFNg+, IL-2+, TNFa+) at different infection stages. I) Kinetics of multifunctional CD4+ T cell response. J) Comparison of multifunctional CD4+ T cell response during the acute phase of infection in infants and young children and adults. Statistical comparisons were conducted with Wilcoxon rank sum test. Solid line indicates median healthy response; dashed line indicates 3x median healthy response.

### Kinetics of T and B cell responses

To further define the adaptive immune response to SARS-CoV-2 infection in infants and young children, we determined the frequency of SARS-CoV-2-specific memory B and T cells in 77 PBMC samples from 34 infants and young children, and 19 adults (**Figure 2a**). We enumerated SARS-CoV-2 Spike-specific memory B cells (MBCs) with two fluorescently labeled Spike probes (S) against the Wuhan and Omicron strain, respectively (**Supp Figure 3a**). As expected, during the first week of infection, S-specific MBCs were few but emerged rapidly ten days after infection (**Figure 2b**).

**Figure 3.**
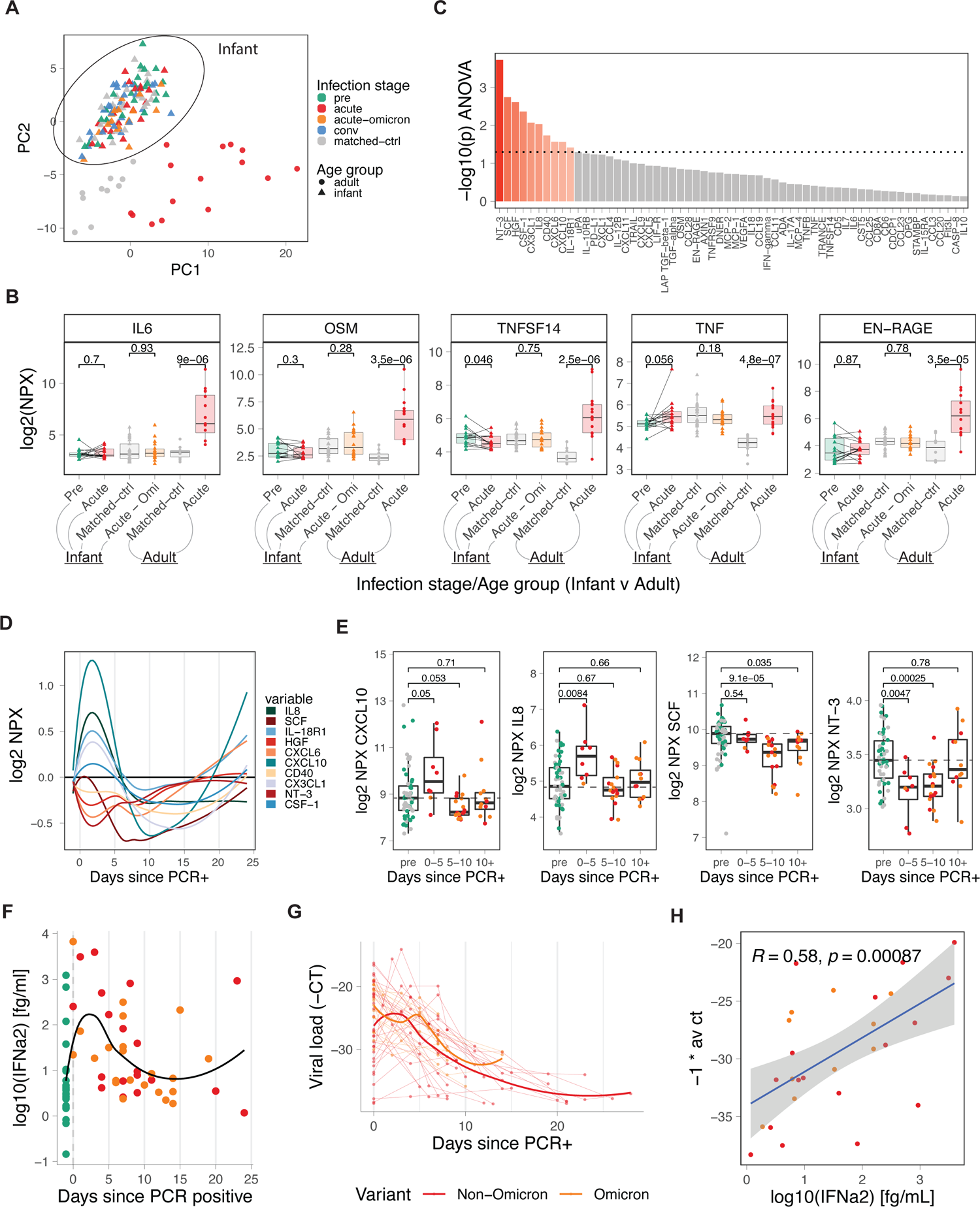
IFN-driven plasma cytokine response to COVID-19 infection in infants and young children. A) PCA analysis of Olink-based plasma cytokines from COVID-19-infected and healthy infants and young children and adults, including matched controls (adult n = 10, infant n = 27), pre-infection (infant n = 27), acute (adult n = 15, infant n = 19), acute-omicron (infant n = 18), and convalescent (n = 30) time points, colored by infections stage. B) Comparison of key inflammatory mediators during COVID-19 infection. Only paired samples are shown for infant pre and acute (n = 14). C) Time-dependent, infection-associated changes in plasma cytokine levels were analyzed using ANOVA. Shown is the p-value for the top 60 analyzed cytokines. Cytokines with an ANOVA p-value < 0.05 are indicated in red. D) Kinetics of indicated plasma cytokines. E) Box plots showing the time-dependent changes in plasma levels of key cytokines. F) Dot plot showing plasma IFN⍺2 levels in infants and young children relative to the first positive COVID-19 test (healthy n = 27, acute n = 19, acute-omicron n = 22). G) Viral load in nasal swabs of COVID-19-infected infants and young children was estimated using RT-qPCR. Line graph showing all −1 * Ct values since the first positive COVID-19 test for each infant (acute n = 32, acute-omicron n = 18. H) Correlation between plasma IFN⍺2 levels and viral load (−1 *Ct). Statistical comparisons were conducted with the Wilcoxon rank sum test (E) and the paired and unpaired t-test (B). Correlation analyses were conducted using Spearman correlation. Lines were fitted using the loess approach.

Importantly, S-specific IgG+ MBCs in infants and young children were maintained for approximately six months after infection but appeared to decline afterward (**Figure 2b, Supp Figure 3b**). This observation differs from studies in adults where MBCs were maintained for more than eight months without signs of decline ^14–16^. Consistent with the serological data (**Figure 1d,e**), in convalescent infants and young children infected with non-Omicron variants, we observed a significantly lower frequency of Omicron S-binding MBCs than that of Wuhan S-binding MBCs (**Figure 2c**). Furthermore, the magnitude of the MBC response in infants during acute infection was lower than in adults (**Figure 2d; Supp Figure 3b,c**). Interestingly, we observed low but detectable frequencies of S-specific class-switched MBCs in several pre-infection infant samples (**Supp Figure 3b**), suggesting the existence of cross-coronavirus-reactive B cells ^19^. To further dissect memory B cell maturation, we sorted S-specific IgG+ MBCs and analyzed their immunoglobulin B cell receptor sequences. Expanded clones with relatively low somatic hypermutation rates (SHM) emerged during acute infection, characteristic of a primary B cell response (**Figure 2e, left**). During convalescence, S-specific MBC clones harbored higher SHM rates (**Figure 2e, right**). Interestingly, the SHM rates in infants and young children were at least comparable, if not higher, than in adults (**Figure 2f**). To evaluate the maturation of the B cell response in each individual infant, we calculated the mean SHM rate of all *IGHV* genes isolated from each infant. In agreement with the observation at the single-cell level, the SHM rate increased in individuals over time (**Figure 2g**), suggesting continuous B cell evolution over the course of months, as has been shown in adults ^20^.

With respect to T cells, we observed an increase of multifunctional Th1-type CD4+ T cells (IL-2, IFN-γ, TNFα triple-positive) in infants and young children post infection (**Figure 4h**) while single-positive CD4+ T cell responses did not exceed the LOD threshold (**Supp Figure 3d)**. Multifunctional CD4+ T cells were induced during acute infection, peaked during the first three months of convalescence, and decayed during late convalescence with relatively low frequencies around 200 days post-PCR+ (**Figure 2h,i, Supp Figure 3e**). Notably, the magnitude of WT and Omicron responses was similar in multifunctional CD4+ T cells (**Supp Figure 3f**). Compared to adults, infant multifunctional CD4 T cell responses were reduced by roughly two orders of magnitude (**Figure 2j**).

**Figure 4.**
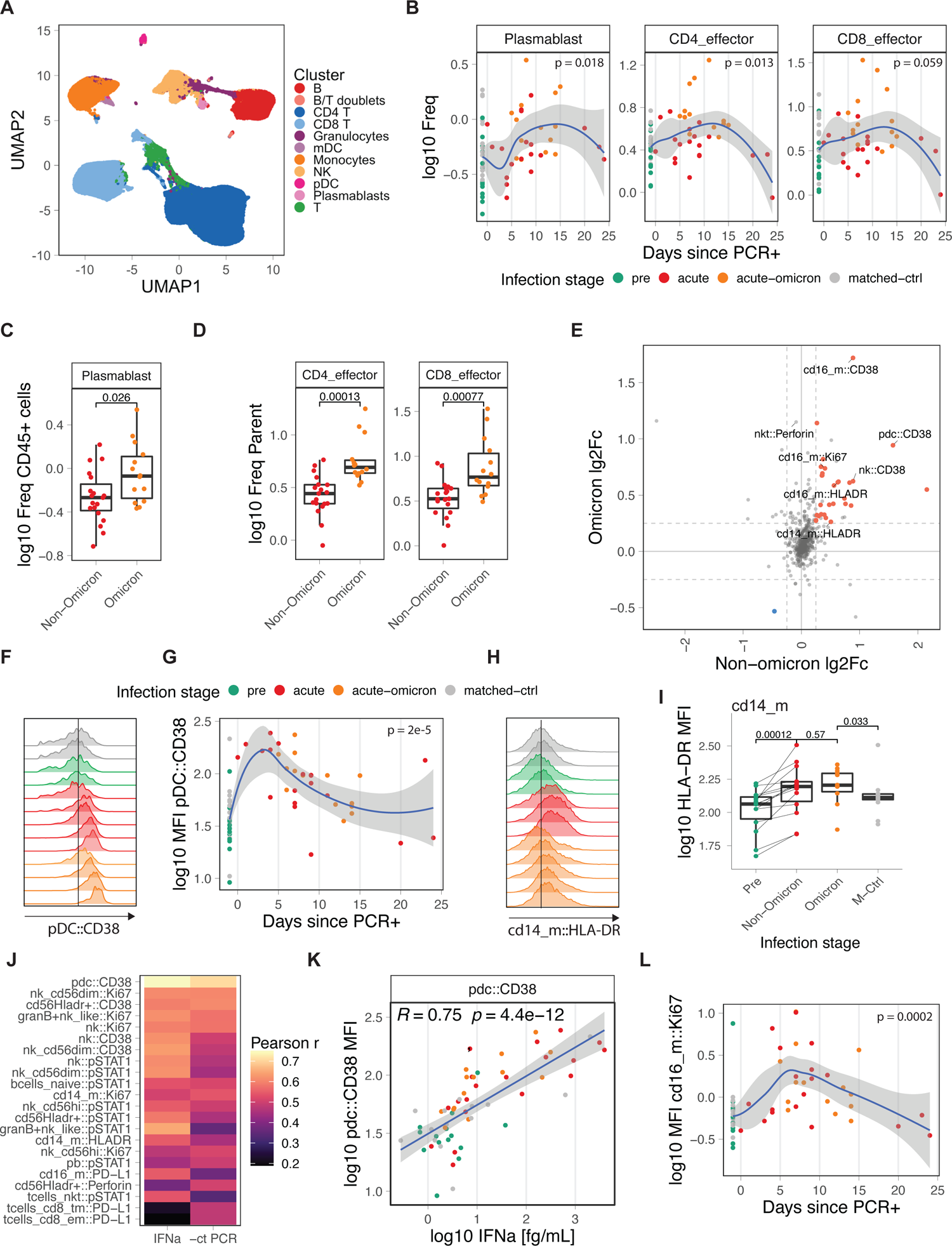
Innate immune cell activation during COVID-19 infection. A) UMAP overview of cell clusters identified by CyTOF (n: pre=14, acute=19, acute-omicron=14, conv=14, matched-ctrl=14). B) Frequency of plasmablasts and effector T cells as a proportion of total CD45+ and total T cells, respectively. C, D) Comparison of plasmablast and effector T cell frequencies in infants and young children infected with Non-Omicron and Omicron variants. E) Scatter plot showing the average log fold-change of marker expression levels in healthy and infected samples for Non-Omicron and Omicron variants. Markers that are significantly changed in both conditions are colored. F) Histogram showing the distribution of CD38 expression in pDCs in representative samples. G) Kinetics of CD38 expression in pDCs. H) Histogram showing the distribution of HLA-DR expression in classical monocytes (CD14_m) in representative samples. I) Box plot comparing HLA-DR expression in classical monocytes (CD14_m). J) Heatmap showing Pearson r for correlations between CyTOF marker expression and plasma IFN⍺2 levels or viral load (−1 * Ct). K) Scatter graph plotting plasma IFN⍺2 levels against CD38 expression in pDCs. L) Kinetics of Ki67 expression in non-classical monocytes (cd16_m). Statistical comparisons were conducted with the Wilcoxon rank sum test. Correlation analyses were conducted using Pearson correlation.

### Kinetics of the plasma cytokine responses

Previous studies have highlighted a dysregulated innate immune system, including the induction of inflammatory plasma cytokine responses, during COVID-19, which was associated with a severe course of disease ^21–23^. To determine plasma cytokine levels during SARS-CoV-2 infection in infants and young children, we used Olink technology and measured the concentration of 96 analytes in 146 plasma samples from 77 infants and young children and 25 adults ^21^. We used Principal Component Analysis (PCA) to determine differences at a global level (**Figure 3a**). Plasma samples from infants and young children, and adults were separated by PC2, while PC1 separated infected and healthy samples among adults (**Figure 3a)**. In contrast, infant samples did not separate based on disease status, indicating profound differences in the cytokine plasma response between infants, and adults (**Figure 3a)** but only minor changes in plasma cytokine levels during infant COVID-19 infection ^10^. Indeed, the expression of proinflammatory cytokines, such as IL6, OSM, TNFSF14, and EN-RAGE, that we and others previously described to be highly elevated in adults during COVID-19 infection ^21–, 23^ were unchanged in infants and young children (**Figure 3b**). Of note, in a previous study, older children (median age 5 years) with predominantly mild disease were shown to upregulate inflammatory markers, such as IL-6, similar to adults ^6^. In contrast, in our cohort, IL-6 expression in infants and young children was still reduced when compared to mild and moderate adult samples only (**Supp Figure 4a**). Instead, we identified a distinct cytokine response in infants and young children during the first weeks and months of life that is characterized by alterations in NT-3, SCF, HGF, CSF-1, CX3CL1, CCL4, CD40, IL8, CXCL10, IL-18R1, CXCL5, CXCL6, and IL-10RB (**Figure 3c**). Longitudinal analysis revealed a transient upregulation for a subset of chemokines (CXCL10, IL8, IL-18R1, CSF-1, CX3CL1) during days zero to five, followed by a transient reduction starting around day five after turning PCR+ (**Figure 3d,e**).

Furthermore, we measured the concentration of IFN-α2 in plasma using a highly sensitive electrochemiluminescence assay. SARS-CoV-2 infection induced robust but transient levels of plasma IFN⍺2 during the first five to ten days after testing positive (**Figure 3f**), similar to what was previously observed in adults ^21^. Notably, IFN⍺2 levels were comparable between infants and young children infected with Omicron and Non-Omicron variants (**Supp Figure 4b**). Since type I IFNs are induced by viral infections, we measured viral loads using PCR in nasal swabs. Kinetic analysis revealed a peak viral load around day zero to five after initial positive testing, with most tests turning negative again after day 20 (**Figure 3g**). Strikingly, there was a strong correlation between plasma IFN⍺2 levels and viral loads (**Figure 3h**). In line with previous studies, infants and young children, and adults mounted comparable IFN⍺2 responses (**Supp Figure 4c**) ^6^.

### Analysis of peripheral blood leukocytes by mass cytometry

Given the observed differences in adult and infant plasma cytokine and chemokine levels and the rapid induction of type I IFNs, we wondered whether infants and young children also display a qualitatively distinct cellular immune response to SARS-CoV-2. Thus, we profiled 75 PBMC samples from 47 infants and young children with cytometry by time of flight (CyTOF) using a panel of 39 antibodies against cell surface markers and intracellular signaling molecules (**DataS3**). Unsupervised clustering analysis revealed 11 cell types (**Figure 4a)**, which we further subtyped by manual gating (**DataS3, Supp Figure 5a**). In line with our analysis of antigen-specific T and B cell immunity (**Figure 2**), frequencies of plasmablasts and CD4 T cells were increased after infection (**Figure 4b**) as previously observed in adults ^21^. Interestingly, plasmablasts and effector T cell frequencies were higher in infants and young children infected with the Omicron variant compared to WT or Delta variants (**Figure 4c,d**). In contrast to the antigen-specific T cell analysis (**Figure 2h)**, we did observe a trend towards elevated levels of CD8 effector cells during the acute phase using CyTOF (**Figure 4b)**. The frequencies of plasmablasts and effector CD4 and CD8 T cells were associated with plasma IFN⍺2 levels (**Supp Figure 5b).**

**Figure 5.**
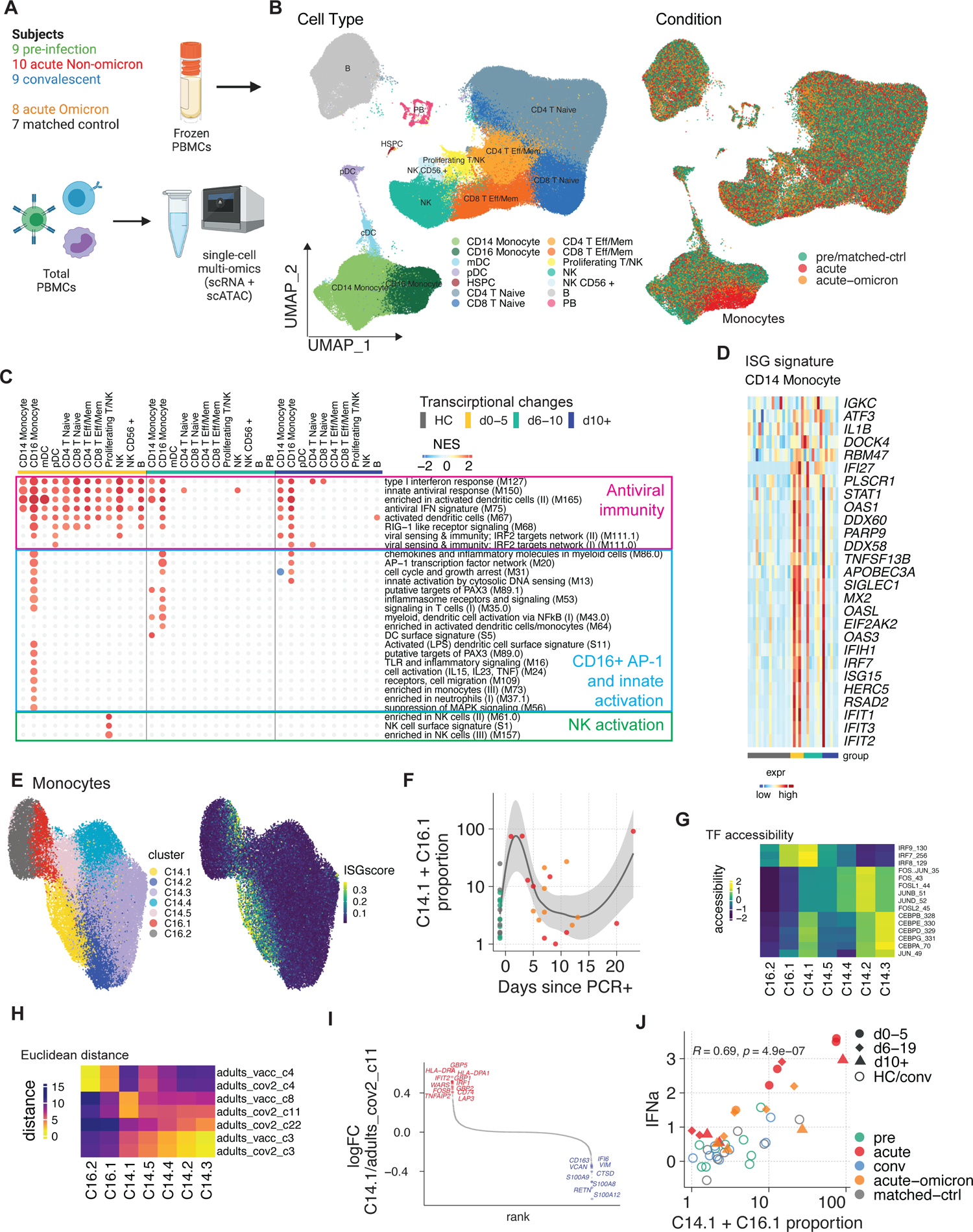
Single-cell multi-omics analysis of immunity to COVID-19 infection in infants and young children. A) Cartoon of the conducted experiment. B) UMAP representation of PBMCs from all analyzed samples, colored by cell type (left) and infection stage (right; convalescent samples not shown). C) Pairwise comparison of genes from healthy (n = 16) and COVID-19–infected infants and young children at different times during acute infection (D0-5: n = 5, D5-10: n=7, D10+: n=6) was conducted for each cluster. DEGs were analyzed for the enrichment of BTMs. Ring plot shows an abridged representation of enriched pathways in each cluster. Size indicates the number of samples with enrichment; colors indicate the normalized enrichment score. Full ring plot in Supp Figure 6a. D) Heatmap showing expression of ISGs enriched in CD14+ monocytes in C) (magenta box). E) UMAP representation of monocyte subclustering analysis. F) Kinetics of CD14.1 and C16.1 monocyte subsets. G) Chromatin accessibility for selected TFs in different monocyte subsets. H) Integrated analysis of monocyte clusters from this study and from adult COVID-19 patients ^21^ and adult subjects immunized with the COVID-19 vaccine ^30^. Shown is the Euclidean distance between infant and adult monocyte subsets. I) DEGs determined between infant C14.1 and adult COVID-19-infection C11 monocyte clusters are plotted and ranked by fold change. J) Spearman’s Rho correlation analysis between plasma IFN⍺2 levels and fraction of interferon-experienced monocytes (bottom).

With respect to the innate immune system, cells from infection with both Omicron and pre-Omicron variants displayed elevated levels of activation, effector, and proliferation markers, including CD38, HLA-DR, perforin, and Ki67, on pDCs, NK cell subsets, and monocytes (**Figure 4e, Supp Figure 5c)**. In adults, we previously observed functional impairment of innate immune cells during COVID-19, which was characterized by reduced frequencies of pDCs in PBMCs, lower expression of pS6 in pDCs, a downstream mediator of mTOR signaling, and reduced levels of HLA-DR expression in monocytes and myeloid dendritic cells ^21, 24^. In infants and young children, we did not observe such impairment (**Supp Figure 5d,e**). Instead, CD38 and HLA-DR expression levels on pDCs and mDCs and classical monocytes (cd14_m), respectively, peaked early during infection (**Figure 4f-i, Supp Figure 5e)** and strongly correlated with IFN⍺2 plasma levels and viral load (**Figure 4j,k**). Of note, a recent study in individuals infected with influenza reported that CD38 upregulation is associated with IFN⍺ production by pDCs *in-vitro* ^25^. Phosphorylated STAT1 levels, a direct downstream mediator of IFN⍺2 signaling and an essential regulator of antiviral immunity ^26^ correlated with IFN⍺2 levels in various cell types (**Figure 4j**), suggesting that IFN⍺ could be a potential driver of the observed innate activation ^27, 28^. Finally, we observed the concerted upregulation of a set of activation markers (HLA-DR, CD38, Ki67) on non-classical monocytes (cd16_m) (**Figure 4e,l**). In contrast to classical monocytes, non-classical monocyte activation kinetics were delayed with Ki67 expression, for instance, peaking during the later phase of infection on days five to 15, indicating a distinct and prolonged response program in these cells.

### Single-cell transcriptional landscape

To gain further mechanistic insights into the activation state of individual immune cells, we used a single-cell multi-omics approach and determined paired gene expression and chromatin accessibility profiles in individual cells in 43 samples from before, during, and after infection with SARS-CoV-2 (**Figure 5a**). Using dimensionality reduction and clustering approaches, we constructed a map of the transcriptomic and epigenomic landscape and identified all major immune cell types (**Figure 5b**). Global analysis of differentially expressed genes demonstrated the induction of antiviral and interferon-related pathways in many cell types on days zero to five after PCR+ (**Figure 5c, Supp Figure 6a**). While this signature waned rapidly in most cells, monocytes expressed elevated levels of antiviral and interferon-related genes for more than ten days post-PCR+ (**Figure 5c**). Analysis of the driver genes revealed enrichment of interferon-stimulated genes (ISG), including STAT1, MX2, IRF7, and multiple members of the IFIT and OAS families (**Figure 5d**).

**Figure 6.**
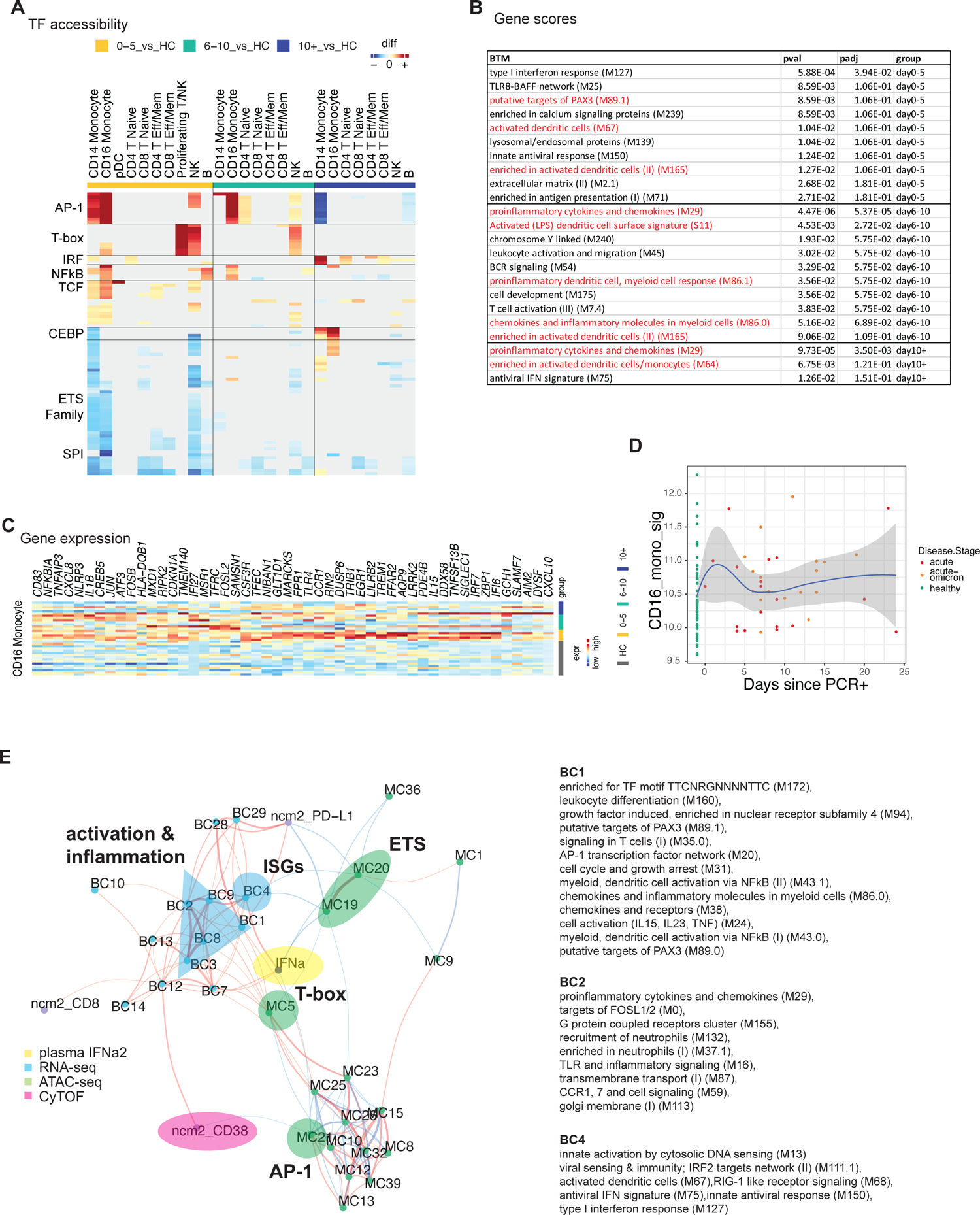
Single-cell multi-omics analysis of CD16+ monocyte activation. A) Pairwise comparison of TF motif accessibility was conducted for each cluster (healthy: n=16; D0-5: n=5, D5-10: n=7, D10+: n=6). Color indicates the difference in TF accessibility; non-significant changes (FDR>=0.001 or changed in less than three subjects) are grey. B) Table showing enrichment of BTMs in differentially accessible gene scores in CD16+ monocytes at indicated time points. C) Heatmap showing expression of inflammation and AP-1-related genes enriched in CD16+ monocytes in Figure 5c (blue box). D) Kinetics of gene signature from (C) using bulk transcriptomics data (healthy: n=53, acute: n=19, acute-omicron: n=18). E) Integrated network analysis of plasma IFN⍺2 levels, BTM-based gene expression, TF motif accessibility, and CyTOF protein marker expression. Both line color and thickness indicate Spearman’s rank correlation coefficient.

Next, we asked how the observed acute-phase ISG response in monocytes is regulated at the single-cell level. Subclustering analysis revealed two subsets of CD14+ and CD16+ monocytes (C14.1, C16.1) that emerge early during the acute phase and are characterized by elevated ISG expression (**Figure 5e, f)**. Using the chromatin accessibility data from our multi-omics dataset, we identified regulatory features associated with the observed monocyte subsets: C14.1 monocytes were characterized by high chromatin accessibility at IRF loci and reduced accessibility at AP-1 loci (**Figure 5g**). These data suggest that a subset of ISG-expressing cells with elevated IRF accessibility drives the observed acute antiviral response in monocytes on a single-cell level. Previously, we identified a subset of monocytes with elevated IRF and reduced AP-1 accessibility after vaccination with an AS03-adjuvanted influenza vaccine ^29^, and we showed that an ISG-expressing monocyte subset emerges rapidly after secondary immunization with the Pfizer/BioNTech COVID-19 vaccine, BNT162b2 ^30^. To determine whether C14.1 and C16.1 are unique monocyte subsets that only emerge in infants and young children or whether they are present in adults as well, we integrated scRNA-seq data from the present study with data from our previous studies on COVID-19 vaccination ^30^ and infection ^21^ in adults and constructed a joined monocyte landscape (**Supp Figure 6b**). In this joint space, infant C14.1 cells overlapped with both adult ISG-expressing cells (C8, adult vaccine study) and a cluster of IFN-activated monocytes (C11, adult COVID-19 study), as demonstrated by UMAP and proximity analysis using Euclidean distance (**Figure 5h, Supp Figure 6b**). Direct comparison of gene expression signatures revealed that infant C14.1 monocytes express elevated levels of ISGs (MX1,2, ISG15, IFI1, IFI44) and antigen presentation markers (HLA-DR) and reduced levels of inflammatory and monocyte activation markers (GBP5, S100A8, FCGR1A) (**Figure 5i, Supp Figure 6c**). Thus, the transcriptional signature of C14.1 in infants and young children resembled the vaccine-induced monocyte cluster more than that of the monocytes observed in COVID-19+ adults. Correlation analysis demonstrated a strong association between ISG expression and plasma IFN⍺2 levels in all immune cell subsets (**Supp Figure 6d**). Monocytes stood out as the cell type with the strongest correlation and the highest expression of ISGs, indicating that these cells are most sensitive to IFN signaling. In line with these findings, we observed a strong correlation between IFN⍺2 and ISG-expressing monocyte subsets (C14.1, C16.1) on a single-cell level (**Figure 5j**).

### Single-cell epigenetic landscape

Next, we determined global changes in chromatin accessibility during infection. TF motif-based analysis revealed extensive chromatin rearrangements in monocytes and NK cells early during infection (**Figure 6a**). Especially, CD16+ monocytes displayed increased chromatin accessibility at gene loci associated with inflammation and immune activation, including AP-1, NFⲕB, and T-box family members (**Figure 6a**). In line with our single-cell data on monocyte heterogeneity, global IRF accessibility was increased in monocytes after infection (**Figure 6a**). Next, we calculated gene score values, aggregating the chromatin accessibility at multiple loci associated with a specific gene^31^. Differential gene score analysis followed by overrepresentation analysis demonstrated enrichment of inflammatory and cytokine-related BTM gene modules in differentially accessible genes of CD16+ monocytes (**Figure 6b**), especially during the later phase of infection. Similarly, gene expression data demonstrated a prolonged activation signature with AP-1, PAX-3, and other inflammation-related BTMs in these cells (**Figure 5c**) that was driven by monocyte activation and inflammation-related genes, including AP-1 TF genes (CD83, NFKBIA, NLRP3, FOSB, JUN, FOSL2, TLR4) (**Figure 6c).** CD16+ monocytes also upregulated IL-8 (CXCL8) and CXCL10 expression, suggesting that they might be a cellular source of the observed plasma chemokine response (**Figure 3d,e)**. While many of these genes were already upregulated early after infection (**Figure 6c**), single-cell and bulk gene expression analyses indicated a subset of primarily inflammatory genes (CD83, NFKBIA, CXCL8, IL1B, NLRP3) that were elevated in some samples also during the later phase of infection (**Figure 6c,d**). In addition to CD16+ monocyte activation, NK cells showed an increase in accessibility at T-box, and AP-1 TF loci and differential gene expression analysis demonstrated the enrichment of BTMs and genes associated with NK cell function early after infection (**Figure 5c, Supp Figure 6e**).

Recent studies by us and others have shown that immunological events, including vaccines and infections, can have a lasting impact on the epigenomic landscape of the innate immune system ^29, 32–34^. To assess whether COVID-19 infection during early life also had a lasting impact on TF accessibility, we determined changes in chromatin accessibility in convalescent samples. To account for potential age-related changes in the epigenetic landscape, we compared the convalescent samples with samples from age- and sex-matched healthy controls. Using this approach, we observed a reduction in CEBP and AP-1 accessibility in CD14+ monocytes of convalescent infants and young children (**Supp Figure 7a**). Importantly, sample-level analysis demonstrated profound interindividual heterogeneity in chromatin accessibility levels, with only a subset of infants and young children showing a reduction in CEBP and AP-1 accessibility (**Supp Figure 7b)**.

**Figure 7.**
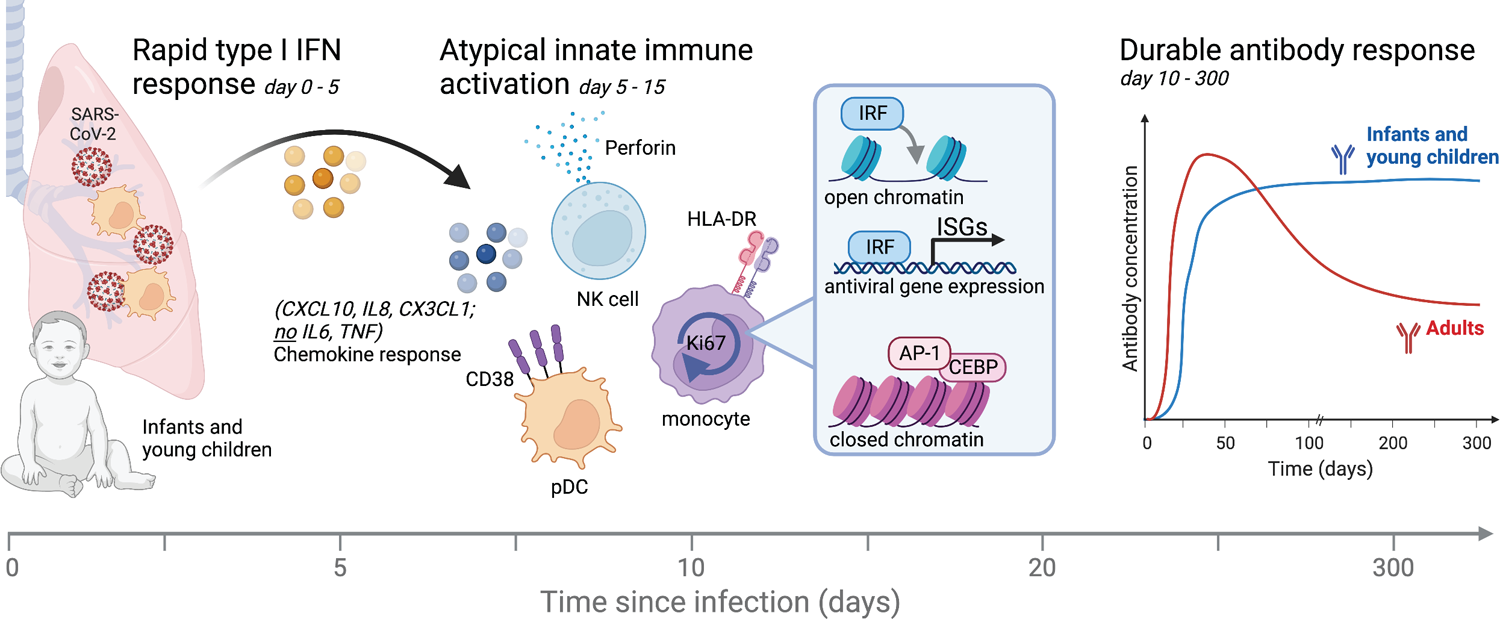
The dynamics of innate and adaptive immunity to a primary infection during the first weeks and months of life. Cartoon summary of major findings.

Finally, to better understand the relationship between the observed ISG response and the distinct innate activation and inflammation profiles in CD16+ monocytes, we integrated our sequencing-based single cell multi-omics analysis with other measurements from this study and constructed a multi-omics network (**Figure 6e**). Our network integrated data on IFN⍺2 (**Figure 3f)**, TF accessibility (**Figure 6a**), gene expression (**Figure 5c**), and CyTOF-based immune profiles (**Figure 4**). In line with our single-cell data, plasma IFN⍺2 levels were positively correlated with the expression of ISGs (BC4) and cell adhesion genes (BC7, BC12) (**Figure 6e**). However, no strong association with TF accessibility was observed (**Figure 6e**), indicating that gene loci associated with ISGs are already in an open chromatin confirmation at baseline. Indeed, analysis of gene tracks at key antiviral regions confirmed the presence of open chromatin regions at the promoter and distal loci before infection (**Supp Figure 7c**). CD16+ monocytes upregulated an additional gene program during acute infection characterized by inflammation, innate activation, PAX3, and AP-1 BTMs (**Figure 5c**). Interestingly, IFN⍺2 was not associated with those genes (BC1) (**Figure 6e**). In fact, IFN⍺2 levels were negatively associated with AP-1 (MC2) TF accessibility (**Figure 6e**), suggesting the induction of a distinct and IFN⍺-independent inflammation program in CD16+ monocytes. To our knowledge, this innate activation and inflammation state of CD16+ monocytes is unique to infants and young children and was not observed in older children or adults ^6, 21^. Together, these data demonstrate that CD16+ monocytes, in addition to the induction of ISGs, upregulate a second, distinct, and seemingly IFN-independent epigenomic and transcriptional profile characterized by genes and TFs associated with innate immune activation and inflammation.

## Discussion

Here, we used a multi-omics approach to study immunity to SARS-CoV-2 infection in infants and young children. Previous reports suggest infants develop attenuated adaptive immune responses compared to adults ^2, 3^. In our study, we observed robust and durable antibody responses against SARS-CoV-2. In contrast, prior studies in adults have shown a decay of antibody responses after COVID-19 infection with a half-life of approximately 120 days ^14, 16^. The mechanisms underlying sustained antibody titers in infants are currently unknown and, given their importance for long-term vaccine protection, demand further investigation. A possible explanation could be that the bone marrow of infants, who have experienced fewer antigenic stimulations than adults, provides a larger number of supportive niches to sustain long-term plasma cell survival^35^. Despite these results, our data also suggest a potential vulnerability against emerging variants and variants of concern (VOC): 1) serum neutralization titers against Omicron variants (BA.1, BA.2, BA.3, BA.4/5) were diminished in infants and young children infected with wildtype or Delta variants and vice versa, as also shown by others ^36^; 2) memory B cell responses, an essential source of affinity matured, cross-specific antibodies, were of limited duration and magnitude; 3) memory T cell responses, which provide broad immunity that is less susceptible to mutational changes and VOC, were reduced compared to adults.

With respect to autoantibodies, we observed an increase in antibody levels against multiple autoantigens, including proteins associated with SLE, myositis (SRP54), autoimmune overlap syndromes (MI-2, Jo1), and scleroderma. Importantly, we did not observe an increase in anti-IFN⍺ autoantibodies in the serum of COVID-infected infants and young children relative to that in healthy control infants. Importantly, age-matched control samples of infants and young children without COVID-19 infection showed similar autoantibody levels as convalescent samples suggesting that an increase in autoantibodies after birth might be a common feature of the developing infant immune system, potentially caused by exposure to other infections. However, it should be emphasized that a potential caveat to this is the transfer of maternal immunoglobulins from mothers to infants before birth could result in circulating antibodies in the infant during the initial months of life.

With respect to innate immunity, we identified a non-canonical state of inflammation characterized by upregulation of activation markers on blood innate cells, and a plasma cytokine profile distinct from that seen in adults, with no inflammatory cytokines, but an early and transient accumulation of chemokines, and type I IFN. While previous studies identified high levels of TNFa, IL-6, OSM, EN-RAGE, and other inflammatory mediators in older children ^6^ and adults ^21–23^ during acute COVID-19, we could not detect any of these cytokines in infants and young children. In contrast, acute plasma samples from infants and young children displayed high levels of IFN⍺2, as well as CXCL10, IL-8, and IL−18R1. Our analysis using CyTOF and single-cell multi-omics further revealed rapid activation of innate immune cells displaying elevated levels of activation markers (CD38, HLA-DR), phosphorylated STAT1, and ISGs. Importantly, in monocytes, this antiviral response was driven by a subset of ISG-expressing cells with elevated IRF accessibility. These results are in striking contrast to results in adults, especially in patients with severe COVID-19, showing major defects in myeloid cells and plasmacytoid dendritic cells during acute infection ^21, 24, 37^. CD16+ monocytes, in addition to the antiviral response, displayed an AP-1 and Pax3-driven inflammatory program independent of IFN⍺. CD16+ monocytes have been implicated with RNA virus infection in previous studies, including with Dengue ^38^ and Zika ^39^. In COVID-19, several studies in adults reported diminished levels of non-classical monocytes ^24, 37^. Together, the lack of inflammatory markers and the rapid induction of innate antiviral immunity might contribute to the mild course of disease observed in infants and young children by containing viral replication early on and preventing the development of severe symptoms. Multiple reasons for the altered innate immune response in infants and young children are conceivable, including breastfeeding, which has been shown to have anti-inflammatory effects ^40, 41^ whilst enhancing type I IFN production ^42^. Furthermore, previous in-vitro studies have demonstrated a defect in TLR-induced inflammatory cytokine production in neonates ^43, 44^. It is thus conceivable that similar mechanisms also prevent the production of inflammatory cytokines upon TLR-mediated SARS-CoV-2 recognition ^45^. A caveat in our analysis is that adult samples are derived from mild, moderate, and severe COVID-19 patients, while infants had predominantly mild disease. Interestingly, even when compared to mild adult cases only, infants displayed a reduced inflammatory plasma cytokine profile (**Supp Figure 5a**). Nevertheless, even if the non-canonical innate response of infants to SARS-CoV-2 could simply reflect the response observed in adults with mild disease, this still begs the important question of why a much greater proportion of infants compared to adults develop such non-canonical innate responses and mild disease.

Previous studies have highlighted the role of vaccine- and infection-induced persistent epigenomic changes in shaping immune responses ^12, 29, 32^. An important question is how COVID-19 infection impacts the epigenomic immune cell landscape in infants and young children. In our study, we observed enhanced accessibility of chromatic loci targeted by interferon regulatory factors (IRFs) and reduced accessibility of AP-1 targeted loci, as well as traces of epigenetic imprinting in monocytes, during convalescence. In particular, convalescent infants and young children displayed reduced levels of CEBP and AP-1 accessibility. Importantly, these epigenetic and cellular changes were highly heterogeneous between individuals. These findings contrast with previous adult studies demonstrating concerted epigenomic and functional changes in monocytes ^29, 32^. Multiple factors could explain this discrepancy: infants and young children experience immunological events (infections, vaccinations) at a high rate. Repeated activation of the immune system between sampling time points could have blurred the epigenomic imprint of COVID-19. In addition, COVID-19 infections in our cohort were mild, possibly leading to faint epigenomic imprints with limited duration. Furthermore, the impact of immune system development and maturation could further alter COVID-19-induced epigenomic changes. Our findings suggest that epigenetic imprints in the infant immune system could be relatively short-lived and more heterogeneous than in adults.

To the best of our knowledge, our study is the first to perform a multi-omics, longitudinal systems immunology analysis of infants. Our comprehensive multi-omics profiling provides insights into perturbations induced by a pathogen in a relatively immature immune system. It has the potential to serve as a reference dataset for future multi-cohort studies that integrate data across multiple cohorts ^46, 47^ to further investigate changes in the immune response to pathogens ^46, 47^.

In summary, our findings provide insights into the dynamics of human immunity to an infection in early life and reveal a surprisingly robust and durable antibody response in the face of an atypical innate response to that observed in adults, characterized by a lack of inflammatory cytokines, but an early and transient type I IFN response and plasma chemokines (**Figure 7**). This apparent disconnect between the adaptive immune response and the pro-inflammatory cytokine response, suggests that a non-canonical pathway of innate activation might drive persistent humoral immunity in this special population in early life. This raises the prospect of devising vaccine adjuvants that target such non-canonical pathways of innate activation to stimulate persistent antibody responses, without the collateral immunopathology that often results from unwanted inflammation.

## Supporting information

DataS1 - Clinical tables

DataS2 - Autoantibody antigen list

DataS3 - Flow and CyTOF antibody tables and gating

## Data Availability

The single-cell multi-omics and bulk transcriptomics data are currently being submitted to the Gene Expression Omnibus (GEO) and will be publicly available at the time of publication.

## Acknowledgments

We thank all study participants and the Cincinnati Children’s Hospital Medical Center staff and faculty who conducted the clinical work with pediatric samples and isolated and banked specimens, especially Monica M McNeal and Brendon White. We are grateful to the Hope Clinic and Emory Children Center staff and faculty, especially Nadine Rouphael, and the Stanford Medical Center for collecting adult samples. We thank the Human Immune Monitoring Center (HIMC), the Parker Institute for Cancer Immunotherapy (PICI), and the Stanford FACS facility for maintenance and access to flow cytometers and FACS sorting; Stanford Functional Genomics Facility for technical assistance; Dhananjay Wagh for library preparation; John Coller for data analysis. Cartoons were created with BioRender.com.

## Author contributions

Conceptualization: B.P. and F.W.; Investigation: F.W., Y.F., P.S.A., M.H., L.N., D.J., M.T., M.A., L.B., U.A., S.K., K.C.K., M.B., K.P.; Data curation and analysis: F.W., Y.F., H.Z., P.S.A., K.C.K., E.Y., G.T., T.H., D.B.H., and B.P.; Patient recruitment and clinical data curation: A.R.B., S.S., M.A.S., R.S.C., K.C.N.; A.G. and A.S. provided peptides for T cell assays; Supervision: B.P., M.A.S., P.K., S.Kh., T.T.W., P.J.U., J.W., K.C.N., H.T.M., and S.E.B.; Data visualization: F.W., H.Z., Y.F., K.C.K., and T.H.; Writing: F.W. and B.P.; Funding acquisition: B.P. All the authors read and accepted the manuscript.

## Data and materials availability

The single-cell multi-omics and bulk transcriptomics data are currently being submitted to the Gene Expression Omnibus (GEO) and will be publicly available at the time of publication. Datasets will be made available for reviewers earlier if required.

## Materials and Methods

### Cohort and study design

Specimens for this analysis were collected as part of the ongoing IMPRINT Influenza Cohort, an NIH-funded longitudinal birth cohort of healthy mothers and children in Cincinnati, Ohio. Pregnant women were screened for enrollment at three local delivery hospitals, and mothers were enrolled in the third trimester of pregnancy. Starting at two weeks of life, research coordinators trained mothers to complete twice weekly SMS surveys for their child to report on the presence or absence of symptoms common to respiratory infections. Mothers were also trained to collect midturbinate nasal swabs from themselves and from their children, which were analyzed at an IMPRINT laboratory. Study participants were asked to collect at least one nasal swab each week. An additional swab was submitted if either the mother or the child was symptomatic of respiratory illness. Among children enrolled in this cohort, we included 19 pre-omicron SARS-CoV2 cases and 22 omicron SARS-CoV2 cases. Cases were selected based on the timing of pre-infection, acute and convalescent blood collections, and availability of PBMCs at these time points. A set of 30 children, age- and sex-matched to pre-Omicron child cases, were selected as controls; age-matching was based on the age of the case at the time of convalescence. COVID-19 infections were considered symptomatic if the child experienced any symptom at least once, within −7/21+ days of their first COVID-19 positive swab, as reported on weekly surveys. Illness severity was evaluated by the presence of cough, fever or difficulty breathing, duration of cough and/or fever, and receipt of medical attention. The presence or absence of symptoms on the day of blood collection was determined by responses on the weekly survey completed closest to the blood collection date (−3/+3 days of blood collection). If a weekly survey response was unavailable, responses from the visit questionnaire were used (n=1). Demographic data is shown in **DataS1**.

### SARS-CoV-2 RT-PCR

Nasal swab samples underwent nucleic acid extraction using a custom protocol and the QIAamp 96 Virus Kit on the QIAGEN QIAcube HT instrument. Extracted RNA was run on Applied Biosystems 7500 or 7500 Fast instruments using CDC-developed real-time RT-PCR assays for Influenza ^48, 49^, SARS-CoV-2 ^50, 51^, or Influenza and SARS CoV-2 in the Flu SC2 Multiplex assay ^52, 53^. Nasal swabs were run on the influenza assay from November 2019-March 17, 2020. SARS-CoV-2 PCR was run for nasal swabs collected starting February 1, 2020, and was used in the lab through December 31, 2021. Starting on January 3, 2022, the Flu SC2 assay has been used for SARS-CoV-2 testing. In addition, all infant nasal swabs underwent multiplex detection of 21 respiratory pathogens using the Luminex NxTAG-RPP assay ^54^, including Influenza A and B. All samples positive for Influenza A and/or B underwent additional RT-PCR detection for Influenza A subtype and Influenza B lineage ^48, 49^.

### SARS-CoV-2 sequencing

Nasal swab samples underwent nucleic acid extraction using the QIAGEN Viral RNA Mini Kit (Qiagen, Inc) on the QIAGEN QIAcube Connect instrument following the manufacturer’s recommendations. RNA samples were subjected to direct sequencing using the modified ARTIC3 protocol ((https://artic.network/ncov-2019) with the addition of primer booster sets as implemented in the Qiagen QiaseqDirect protocol. RNA was subjected to random primed cDNA synthesis followed by amplification in two pools of multiplexed primer sets resulting in overlapping amplicons spanning the entire genome. Subsequently, 24 cycles of polymerase chain reaction were utilized to add dual index primers and amplify SARS-Cov-2 amplicons. DNA concentrations were normalized, samples were pooled and then subjected to sequencing to a depth of at least 100,000 reads per sample utilizing paired 150 nucleotide reads on an Illumina NextSeq 500 sequencing machine (Illumina, Inc).

Raw sequence data were demultiplexed and then aligned against the ancestral Wuhan-1 genome (Accession MN908947) ^55^ using bwa-mem ^56^. Samtools commands “sort”, “index”, “view”, and “mpileup” ^57^ were applied sequentially, and the ivar “consensus” command ^58^ was used to output a consensus sequence. Variant identification and lineage calling were performed with the software Phylogenetic Assignment of Named Global Outbreak Lineages (Pangolin) version 4.0.4 ^59^.

### Sample processing

Peripheral blood was collected from study children at two and six weeks of life, every summer and acutely following “events.” An event included receipt of influenza or COVID-19 vaccines or having a nasal swab test positive for influenza or SARS-CoV2; participants completed an additional symptom survey at event visits. Up to 16mL of collected blood was deposited into sodium citrate Mononuclear Cell Preparation tubes (CPT) and promptly delivered to the laboratory for processing. After the initial spin of the CPT tubes, plasma was collected, and up to ten aliquots of plasma at 0.5mL were stored at −65°C or colder. Cells were collected, washed, and counted using the automated Vi-CELL XR Viability Analyzer. Aliquots were made at concentrations of either 2.0×10^6^ or 5.0×10^6^ cells in 1mL of Fetal Bovine Serum (FBS) +10% Dimethyl Sulfoxide (DMSO), depending on the number of cells obtained. Once aliquoted, the cryovials were placed into a Mr. Frosty cooler filled with isopropyl alcohol and placed into a −65°C or colder freezer for 24-72 hours. Samples were then placed into liquid nitrogen storage units for long-term storage.

### Adult Cohort

The samples of the adult participants included in the study were collected in 2020 at the start of the pandemic, prior to the advent of vaccines. All samples were collected at the Hope Clinic of the Emory Vaccine Center or at Stanford University. Healthy controls were asymptomatic adults from whom samples were collected prior to the widespread circulation of SARS-CoV-2 in the community. Patients with COVID-19 were defined according to the original WHO guidance and positive SARS-CoV-2 RT-PCR testing by nasopharyngeal swabs as described in our original study ^21^. Patients with COVID-19 were classified as acute (less than 4 weeks from symptom onset or symptomatic at the time of sample collection) or convalescent (more than 4 weeks since symptom onset and asymptomatic or negative SARS-CoV-2 RT-PCR testing and asymptomatic). The severity of inpatient COVID-19 cases was classified based on the adaptation of the Sixth Revised Trial Version of the Novel Coronavirus Pneumonia Diagnosis and Treatment Guidance. Mild/moderate cases were defined as respiratory symptoms with radiological findings of pneumonia. Severe cases were defined as requiring supplemental oxygen. Critical cases were organ failure necessitating intensive care unit (ICU) care. The study received approval from the appropriate Institutional Review Board at Emory (#00022371) and Stanford (#55689) University. All samples were collected after informed consent. Demographic data is shown in **DataS1**.

### Anti-Spike electrochemiluminescence (ECL) binding ELISA

Anti-Spike IgG titers were measured using V-plex COVID-19 panel 23 from Mesoscale Discovery (Cat #K15567U). The assay was performed as per the manufacturer’s instructions. Briefly, the multi-spot 96 well plates were blocked in 0.15 ml of blocking solution with shaking at 700 rpm at room temperature. After 30 min of incubation, 50 μl of plasma samples serially diluted in antibody diluent solution and serially diluted calibrator solution was added to each plate in the designated wells and incubated at room temperature for 2 h with shaking. Plasma samples were assayed at a 1:100 starting dilution and 6 additional 5-fold serial dilutions. After 2 h of incubation, the plates were washed, 50 μl of Sulfo-tag conjugated anti-IgG was added, and the plates were incubated at room temperature for 1 h. After incubation, the plates were washed, and 0.15 ml of MSD-Gold read buffer was added. The plates were immediately read using the MSD instrument. The Meso scale arbitrary light unit signal was used for calculating the area under curve (AUC) in Prism v.9.4.1.

### Pseudovirus production and neutralization assay

VSV-based GFP/nanoluciferase-expressing SARS-CoV-2 pseudoviruses were produced as described previously ^60^. VSV-ΔG-GFP/nanoluciferase and plasmids encoding spike genes of SARS-CoV-2 Wuhan (SΔ19), Delta (B.1.617), and Omicron (B.1.529) were provided by Dr. Gene S. Tan (J. Craig Venter Institute, La Jolla, CA). To perform the neutralization assay, Vero E6-TMPRSS2-T2A-ACE2 cells (BEI Resources, NIAID; NR-54970) were seeded at a density of 1×10^4^ per well in half area 96-well black opaque plates (Greiner Bio-One) and were grown overnight at 37°C in a 5% CO2 atmosphere. Serum samples were 5-fold serially diluted using the infection medium (DMEM supplemented with 2% FBS and 100 U/mL Penicillin-Streptomycin) in duplicates. Diluted serum samples were then mixed with an equal volume of Wuhan, Delta, or Omicron pseudoviruses, diluted in infection medium at an amount of 200-400 focus-forming units/mL per well, followed by incubation at 37°C for 1 hour. Subsequently, immune complexes were added onto the monolayers of PBS-washed Vero E6-TMPRSS2-T2A-ACE2 cells and incubated at 37°C. At 18 hours post-incubation, supernatants were removed, cells were washed once with PBS, and nanoluciferase enzymatic activities were measured using the Nano-Glo Luciferase Assay System (Promega) and a SpectraMax iD3 multi-mode microplate reader. Percent inhibition values were calculated by subtracting the percent infection from 100. Non-linear curves and IC_50_ values were determined using GraphPad Prism.

### Neutralization assay - Omicron subtyping

Human samples were evaluated in a qualified SARS-CoV-2 pseudovirion neutralization assay (PsVNA) using SARS-CoV-2 WA1/2020 strain and variants. SARS-CoV-2 neutralizing activity measured by PsVNA correlates with PRNT (plaque reduction neutralization test with authentic SARS-CoV-2 virus) in previous studies ^61–63^. Pseudovirions were produced as previously described ^61^. Briefly, human codon-optimized cDNA encoding SARS-CoV-2 spike glycoprotein of the WA1/2020 and variants was synthesized by GenScript and cloned into eukaryotic cell expression vector pcDNA 3.1 between the *BamH*I and *Xho*I sites. Pseudovirions were produced by co-transfection Lenti-X 293T cells with psPAX2(gag/pol), pTrip-luc lentiviral vector, and pcDNA 3.1 SARS-CoV-2-spike-deltaC19, using Lipofectamine 3000. The supernatants were harvested at 48h post-transfection and filtered through 0.45µm membranes, and titrated using 293T-ACE2-TMPRSS2 cells (HEK 293T cells that express ACE2 and TMPRSS2 proteins). Neutralization assays were performed as previously described ^36, 62–65^. For the neutralization assay, 50 µL of SARS-CoV-2 S pseudovirions (counting ∼200,000 relative light units) were pre-incubated with an equal volume of medium containing serial dilutions of samples at room temperature for 1h. Then 50 µL of virus-antibody mixtures were added to 293T-ACE2-TMPRSS2 cells (10^4^ cells/50 μL) in a 96-well plate. The input virus with all SARS-CoV-2 strains used in the current study was the same (2 x 10^5^ relative light units/50 µL/well). After a 3 h incubation, fresh medium was added to the wells. Cells were lysed 24 h later, and luciferase activity was measured using One-Glo luciferase assay system (Promega, Cat# E6130). The assay of each sample was performed in duplicate, and the 50% neutralization titer was calculated using Prism 9 (GraphPad Software). Controls included cells only, virus without any antibody and positive sera.

### Auto-antibody analysis with Bead-based array

We created two different custom, bead-based antigen arrays (**DataS2**) modeled on similar arrays that we previously used to study autoimmune diseases, immunodeficiency disorders, and COVID-19 disease in adults ^11, 66–73^. Antigens were selected based on our published datasets, literature searches that have implicated specific antigens in COVID-19, potential for mechanistic contribution to COVID-19 pathogenesis, and compatibility with bead-based platforms. The autoantigen array contained 60 commercial protein antigens associated with connective tissue diseases and tissues associated with tissue inflammation. The cytokine array comprised 61 proteins including interferons, interleukins, and other cytokines. The arrays were constructed as previously described ^73^. Antigens were coupled to carboxylated magnetic beads (MagPlex-C, Luminex Corp.) such that each antigen was linked to beads with unique barcodes ^68, 70^. Briefly, 8 μg of each antigen or control antibody was diluted in phosphate-buffered saline (PBS) and transferred to 96-well plates. Diluted antigens and control antibodies were conjugated to carboxylated magnetic beads per ID. Beads were distributed into 96-well plates (Greiner BioOne), washed, and re-suspended in phosphate buffer (0.1 M NaH2PO4, pH 6.2) using a 96-well plate washer (Biotek). The bead surface was activated by adding 100 μl of phosphate buffer containing 0.5 mg 1-ethyl-3(3-dimethylamino-propyl)carbodiimide (Pierce) and 0.5 mg N-hydroxysuccinimide (Pierce). After 20 min incubation on a shaker, beads were washed and resuspended in activation buffer (0.05 M 2-N-Morpholino EthaneSulfonic acid, MES, pH 5.0).

Diluted antigens and control antibodies were incubated with beads for 2 h at room temperature. Beads were washed three times in 100 μl PBS-Tween, re-suspended in 60 μl storage buffer (Blocking reagent for Enzyme-Linked Immunosorbent Assay, ELISA, Roche) and stored in plates at 4 °C. Immobilization of some antigens and control antibodies on the correct bead IDs was confirmed by analysis using commercially available monoclonal antibodies or antibodies specific for epitope tags. In addition, prototype human plasma samples derived from participants with autoimmune diseases with known reactivity patterns (from ImmunoVision, Stanford Autoimmune Diseases Biobank, and OMRF) were used for validation. Plasma samples were tested at 1:100 dilution in 0.05% PBS-Tween supplemented with 1% (w/v) bovine serum albumin (BSA) and transferred into 96-well plates in a randomized layout. The bead array was distributed into a 384-well plate (Greiner BioOne) by transfer of 5 μl bead array per well. 45 μl of the 1:100 diluted sera were transferred into the 384-well plate containing the bead array.

Samples were incubated for 60 min on a shaker at room temperature and then stored at 4 °C overnight. Beads were washed with 3 × 60 μl PBS-Tween on a plate washer (EL406, Biotek) and 50 μl of 1:500 diluted R-phycoerythrin (R-PE) conjugated Fcγ-specific goat anti-human IgG F(ab’)2 fragment (Jackson ImmunoResearch, Cat # 109-116-098) was added to the 384-well plate for detection of bound human IgG. After incubation with the secondary antibody for 30 min, the plate was washed with 3 × 60 μl PBS-Tween and re-suspended in 60 μl PBS-Tween prior to analysis using a FlexMap3DTM instrument (Luminex Corp.) and Luminex xPONENT® version 4.2 software. A minimum of 50 events per bead ID were counted. Binding events were displayed as Mean Fluorescence Intensity (MFI). To ensure reproducibility and rigor, all samples were run in duplicate in each experiment. Prototype autoimmune sera as described above, were used as positive controls. All data analyses and figure generation were performed using Python, R and R studio. For normalization, average MFI values for “bare bead” IDs were subtracted from average MFI values for antigen-conjugated bead IDs. The average MFI for each antigen was calculated using samples from healthy infants who have not had COVID and have not tested positive for another upper respiratory infection in the last 7 days. Antibodies were considered “positive” if MFI was >5 SD above the average MFI and >3000 units, which is a threshold that we have used previously for publication and is more stringent than commonly published in related literature ^11, 73^.

### Spike-specific memory B cell staining

Cryopreserved PBMCs were thawed and washed twice with 10 mL of FACS buffer (1 x PBS containing 2% FBS and 1 mM EDTA) and resuspended in 100 uL of 1x PBS containing Zombie UV live/dead dye at 1:200 dilution (BioLegend, 423108) and incubate at room temperature for 15 minutes. Following washing, cells were incubated with an antibody cocktail for 1 hour protected from light on ice. The following antibodies were used: IgD PE (BD Biosciences, 555779), IgM PerCP-Cy5.5 (BioLegend, 314512), CD20 APC-H7 (BD Biosciences, 560734), CD27 PE-Cy7 (BioLegend, 302838), CD14 PE/Dazzle™ 594 (BioLegend, 301852), CD16 BV605 (BioLegend, 302040), IgG BV650 (BD Biosciences, 740596), CD3 BUV737 (BD Biosciences, 612750) and Alexa Fluor 488-labeled Wuhan spike (SinoBiological, 40589-V27B-B), and BV421 labeled Omicron Spike (SinoBiological™, 40589-V49H3-B). All antibodies were used as the manufacturer’s instruction and the final concentration of each probe was 0.1 ug/ml. Cells were washed twice in FACS buffer and immediately acquired on a BD FACS Aria III for acquisition and FlowJo for analysis.

### Single cell BCR-seq

SARS-CoV-2 spike specific memory B cells gated on singlet CD3-CD14-CD16-CD20+ IgM-IgD-CD27^low/high^ IgG+ spikes+ were single-cell sorted into individual wells of 96-well plates containing 16 μl of lysis buffer per well using a FACS Aria III. The lysis buffer was composed of 20 U RNAse inhibitor (Invitrogen), 5 mM DTT (Invitrogen), 4 ul 5x RT buffer (Invitrogen), 0.0625 ul Igepal (Sigma), 10 ug/ml Carrier RNA (Applied Biosystems). The 96-well plates went through a quick freeze-thaw cycle, and 0.5 ug Oligo(dT)18 (Thermo Scientific), 0.5 mM dNTP mix (Invitrogen), and 200 U Superscript IV (Invitrogen) was added in a total volume of 4 ul followed by thorough mixing and spinning. The reverse transcription was performed as follows: 10 min at 42 °C, 10 min at 23 °C, 20 min at 50 °C, 5 min at 55 °C, 10 min at 80 °C and finally cooling to 4°C. Ig heavy chain and light chain (kappa/lambda) variable gene fragments were amplified by nested PCR (HotStarTaq DNA Polymerase, QIAGEN) using primer cocktails ^74, 75^ at a concentration of 250 nM per primer. The PCR mix consisted of 2.5 ul 10x PCR buffer, 0.5 ul 10 mM dNTP mix (Invitrogen), 0.5 ul 25 mM MgCl2, 5 ul Q-solution, 1 U HotStarTaq, 0.5 ul 5’ and 3’ primers and 2.5 ul of cDNA. Water was added up to a total volume of 25 ul. The 2nd round PCR products was evaluated on 2% agarose gels, purified using QIAquick spin columns (Qiagen) and sequenced using 2nd round PCR reverse primers. The sequences were analyzed using the online IMGT/HighV-QUEST tool. IGHV and IGLV nucleotide sequences were aligned against their closest germlines and the somatic hypermutation rate was calculated based on the IMGT v-identity output. The average mutation rate was calculated by dividing the sum of all somatic hypermutation rates by the number of sequences used for the analysis in each individual. The Change-O toolkit v.1.0.0. and SHazaM R package were used for B cell clonality analysis ^76^.

### T cell stimulation and intracellular cytokine staining assay

Antigen-specific T cell responses were measured using the intracellular cytokine staining assay as previously described ^77^. Live frozen PBMCs were revived, counted, and resuspended at a density of 2 million live cells per ml in complete RPMI (RPMI supplemented with 10% FBS and antibiotics). The cells were rested for 6 h at 37 °C in a CO_2_ incubator. At the end of 6 h, the cells were centrifuged, resuspended at a density of 10 million cells per ml in complete RPMI, and 100 μl of cell suspension containing 1 million cells was added to each well of a 96-well round-bottomed tissue culture plate. Each sample was treated with two or three conditions depending on cell numbers: no stimulation or a peptide pool spanning the Spike protein of the ancestral Wu strain or Omicron BA.1 variant (where cell numbers permitted) in the presence of anti-CD28 (1 μg ml^−1^; clone CD28.2, BD Biosciences) and anti-CD49d (clone 9F10, BD Biosciences), as well as anti-CXCR3 (**DataS3**). The peptide pools were 15-mer peptides with 10-mer overlaps spanning the entire Spike protein sequence of each variant ^78^. Each peptide pool contained 253 peptides and was resuspended in DMSO at a concentration of 1 mg/ml.

PBMCs were stimulated at a final concentration of 1 μg/ml of each peptide in the final reaction with an equimolar amount of DMSO [0.5% (v/v) in 0.2-ml total reaction volume] as a negative control. The samples were incubated at 37°C in CO2 incubators for 2 hours before the addition of brefeldin A (10 μg ml^−1^). The cells were incubated for an additional 4 hours. The cells were washed with PBS and stained with Zombie ultraviolet (UV) fixable viability dye (BioLegend). The cells were washed with PBS containing 5% FBS before adding a surface antibody cocktail (**Supp Table 5**). The cells were stained for 20 min at 4 °C in 100-μl volume. Subsequently, the cells were washed, fixed, and permeabilized with cytofix/cytoperm buffer (BD Biosciences) for 20 min. The permeabilized cells were stained with intracellular cytokine staining antibodies (**Supp Table 5**) for 20 min at room temperature in 1× perm/wash buffer (BD Biosciences). Cells were then washed twice with perm/wash buffer and once with staining buffer before acquisition using the BD Symphony Flow Cytometer and the associated BD FACS Diva software. All flow cytometry data were analyzed using Flowjo software v.10 (BD Bioscience). DMSO background was subtracted from all samples and the positivity threshold was defined as 3x the median of peptide stimulated samples from healthy control infants.

### Quantitation of human IFN-α2a in plasma

Human IFN-α2a was measured using an S-PLEX Human IFN-α2a kit from Mesoscale Discovery (Cat # K151P3S). The assay was performed as per the manufacturer’s instructions. Briefly, the uncoated 96 well plates were washed 3 times with 150 μl per well of 1xMSD wash buffer and coated with 50 μl of coating solution per well with shaking at 700 rpm at room temperature. After 1 hour of coating, the plates were washed, and 25 μl of blocking solution was added to each well, followed by adding 25 μl of neat plasma samples or serially diluted calibrator solution to each plate in the designated wells and incubated at room temperature for 1.5 h with shaking. After calibrator and sample incubation, the plates were washed, and 50 μl of TURBO-BOOST antibody solution was added, and the plates were incubated at room temperature for 1 h. After TURBO-BOOST antibody incubation, the plates were washed and 50 μl of enhance solution was added, and the plates were incubated at room temperature for 30 minutes. After enhance solution incubation, the plates were washed, 50 μl of TURBO-TAG detection solution was added, and the plates were incubated with shaking at 27 °C for 1 hour. After TURBO-TAG dection incubation, the plates were washed gently, 150 μl of MSD-Gold read buffer A was added, and the plates were immediately read using the MSD instrument. The IFN-α concentrations were determined by the calibration curves established by fitting the signals from the calibrators using a 4-parameter logistic model with a 1/Y^2^ weighting in Prism v.9.4.1.

### Olink

Cytokines in plasma were measured using Olink multiplex proximity extension assay (PEA) inflammation panel (Olink proteomics: www.olink.com) according to the manufacturer’s instructions as described before ^79^. The PEA is a dual-recognition immunoassay, in which two matched antibodies labelled with unique DNA oligonucleotides simultaneously bind to a target protein in solution. This brings the two antibodies into proximity, allowing their DNA oligonucleotides to hybridize, serving as template for a DNA polymerase-dependent extension step. This creates a double-stranded DNA ‘barcode’ that is unique for the specific antigen and quantitatively proportional to the initial concentration of target protein. The hybridization and extension are immediately followed by PCR amplification and the amplicon is then finally quantified by microfluidic qPCR using Fluidigm BioMark HD system. Normalized Protein eXpression (NPX) values were used for downstream analysis after initial QC filtering. PCA analysis was conducted with the R package “pcaMethods” ^80^.

### CyTOF analysis of PBMC samples

1 million peripheral blood mononuclear cells (PBMCs) were fixed using 2% PFA for 30 min at RT and washed. Fixed PBMCs were then resuspended in freezing media (10% DMSO + 90% FBS), transferred to cryovials, and stored at −80°C until read for downstream processing and staining. As previously described ^21^, fixed frozen PBMCs were thawed in a 37°C water bath and gently resuspended using 1 mL CSM (PBS supplemented with 2% BSA, 2mM EDTA and 0.1 % sodium azide) and transferred to 15 mL conical tubes containing 9 mL CSM. Samples were washed twice using CSM and counted. Cells were then permeabilized and barcoded using the Cell-ID^TM^ 20-Plex Pd Barcoding Kit (Fluidigm, catalog # 201060). Post-barcoding, cells were washed, pooled into one barcode composite, and counted. The pooled composite was then stained for 30 min at RT with a pre-titrated surface antibody cocktail (**DataS3**). After surface staining, cells were washed twice with CSM and fixed in 4% PFA (freshly prepared paraformaldehyde in PBS) for 10 min at RT. Cells were then washed with CSM and permeabilized with 100% cold MeOH (Sigma), and kept overnight at −80°C. The next day, cells were washed with CSM and counted. Intracellular staining was then performed for 30 min at RT with a pre-titrated intracellular antibody cocktail (**DataS3**), followed by two CSM washes. Finally, cells were stained with iridium-containing DNA intercalator (Fluidigm) for 20 min at RT, washed first with CSM and then with MilliQ water. The washed cells were resuspended in MilliQ water supplemented with 1x EQ four element calibration beads (Fluidigm) and acquired on Helios mass cytometer (Fluidigm). The raw FCS files were normalized and concatenated using the Fluidigm software. The normalized .fcs files were then processed in FlowJo software v10 (BD Biosciences) for debarcoding. Briefly, the normalized .fcs file was used to gate single cells based on DNA content and event length in FlowJo. The single cells were then reimported and debarcoded using Helios software version 7.0.5189. The debarcoded samples were analyzed using FlowJo or R version 1.2.1335 for downstream analysis and visualization.

### CyTOF Analysis

High-dimensional analysis of phospho-CyTOF data was performed using a previously described R-based pipeline ^81^. In brief, the raw .fcs files were imported into R, and the data were transformed to normalize marker intensities using arcsinh with a cofactor of 5. For visualization, another transformation was applied that scales the expression of all values between 0 and 1 using percentiles as the boundary. Cell clustering was performed with 4,000 cells randomly selected from each sample using FlowSOM ^82^ and ConsensusClusterPlus ^83^. The transformed matrix was used as an input for FlowSOM, and cells were separated into 20 clusters. To obtain reproducible results (avoid random start), a seed was set for each clustering. The 20 clusters were manually annotated based on the lineage marker expression and were merged to produce the final clusters. The clusters were visualized in two-dimensional space using UMAP ^84^. In parallel, the data were manually gated to identify 34 immune cell subpopulations that were not well-distinguished in UMAP and used for all quantification purposes.

### Single-cell multi-omics experiments

Single-cell ATAC and RNA-seq libraries were prepared using the Chromium Single Cell Multiome ATAC + Gene Expression platform (10X Genomics, Pleasanton, CA). Briefly, cryopreserved PBMCs were thawed and processed for single nuclei multi-omics analysis according to the manufacturer’s instructions (10x Genomics, CG000365 Rev B). Nuclei were obtained by incubating PBMCs for 3.10 minutes in freshly prepared Lysis buffer and washed and resuspended in chilled diluted nuclei buffer (10x Genomics, 2000153). About 9,000 cells were targeted for each experiment. Prepared nuclei were subjected to transposition of open chromatin regions. Next, transposed nuclei, reverse transcription Master Mix, barcoded Gel Beads, and Partitioning Oil were partitioned into single-cell GEMs (Gel Bead in EMulsions) using the 10X Chromium Controller and Next GEM Chip J. Within each GEM, Gel Beads are dissolved and poly-adenlyated (poly-A) mRNA transcripts are captured by uniquely barcoded poly(dt)VN oligos. Simultaneously, accessible chromatin fragments are captured by a separate oligo containing a spacer, unique barcode, and Illumina P5 adaptor sequence. The GEMs were then incubated in a C1000 Touch Thermal Cycler (BioRad) to produce barcodedDNA from the transposed DNA, and full-length cDNA. GEMs are then broken to release and pool single-cell fractions within each sample. Pooled fractions are purified using silane magnetic beads and subjected to PCR amplification to generate sufficient mass for library construction. Next, P7 and a sample index are added to transposed DNA via PCR to generate ATAC libraries. Finally, cDNA is enzymatically fragmented before the addition of P5, P7, i7 and i5 indexes, and TruSeq Read 2 via End Repair, A-tailing, Adaptor Ligation, and PCR resulting in gene expression libraries. Quantitation of gene expression and ATAC libraries was performed using Bioanalyzer High Sensitivity DNA Analysis (Agilent). Libraries were combined into gene expression and ATAC pools and sequenced on an Illumina NovaSeq 6000 system using the read lengths recommended by 10X: 28bp (read 1), 90bp (read 2), 10bp (i7 index), and 10bp (i5 index) for gene expression libraries and 50bp (read 1), 49bp (read 2), 8bp (i7 index) and 24bp (i5 index) for ATAC libraries. CellRanger v.3.1.0 (10xGenomics) was used to demultiplex raw sequencing data and quantify transcript levels against the 10x Genomics GRCh38 reference v.3.0.0.

### Single-cell gene expression analysis

The single cell RNA-seq data was processed with Seurat (v4.0.5) ^85^. We removed cells with less than 800 or greater than 6,000 detected genes, less than 1,000 or great than 60,000 mRNA reads, or greater than 15% mitochondrial reads. Normalization and feature selection were performed using sctransform ^86^. Clusters were identified with Seurat SNN graph construction on Harmony ^87^-corrected PCA embeddings followed by Louvain community detection algorithm. After identification of major cell types, the clustering process was repeated on each cell type separately to get refined clusters, e.g., monocyte sub-clustering, and to remove doublets that are not identifiable in the first-round clustering of all the cells. Differentially expressed (DE) genes were identified using Wilcoxon Rank Sum test in Seurat. For comparison between two groups (e.g., group A vs. group B), two modes of DE analyses were performed: 1) DEall: all the samples in group A vs. all the samples in group B; 2) DEeach: each sample in group A vs. all the samples in group B. For each mode of DE analysis, genes with p-value ≤ 0.01 were ranked by log2 fold change and used as input in gene-set enrichment analysis (GSEA) analysis implemented in the fgsea R package ^88^. Enrichment was assessed with gene lists in Blood Transcriptomic Modules ^89^. For each comparison between two groups, enriched gene sets were filtered according to the following criteria: 1) the gene set was enriched in DEall comparison (adjust p value ≤ 0.05) 2) the gene set was enriched in at least two DEeach comparisons (adjust p value ≤ 0.05). Only gene sets satisfying both criteria were kept. Selected gene sets shown in the main figures were manually curated to select gene sets relevant to immunology and often enriched in several cell types across multiple DE comparisons.

### The ISG score calculation

The ISG score was calculated as the geometric mean of 33 top differentially expressed genes between acute infection and healthy controls (adjusted p value < 0.05, average log2 fold change >= 0.5) across all cell types in 8 BTM terms that are involved in antiviral interferon response (M165, M75, M150, M127, M67, M68, M111.1, M111.0).

### Monocyte single-cell gene expression integration

The single-cell gene expression data of monocytes from the previous adult COVID-19 infections study ^90^ and the Pfizer vaccine study ^91^ were integrated with monocytes in the current study using the Seurat integration workflow with reciprocal PCA algorithm. The normalized expression data in the integrated assay was then used to calculate the distance between clusters and the ISG score in the integrated data. The Euclidean distance between clusters was calculated using the average expression of the top 20 marker genes in each cluster. The folder change between C14.1, adults_cov2_c11, and adults_vacc_c8 clusters was calculated using limma ^92^.

### Single-cell chromatin accessibility analysis

The single-cell ATAC data was processed with ArchR (v1.0.1) ^93^. The cell type annotations were transferred from the single-cell RNA-seq analysis. Transcription factor (TF) binding motifs were annotated using JASPAR 2016 transcription factor binding database ^94^. The per-cell motif deviations and scores were calculated using chromVAR ^95^. The difference in motif deviations between two groups was tested using the Wilcoxon Rank Sum test. Adjusted p values were computed using Bonferroni correction. Similar to DE gene analysis, differential accessibility motifs between two groups (e.g., group A and group B) are defined according to the following criteria: 1) the TF motif deviations are significantly different between all cells in group A and group B (adjust p value ≤ 0.001 for comparisons between acute infection and healthy controls, and 0.0001 for comparisons between convalescent and healthy controls). 2) when comparing cells in each sample in group A vs. all cells in group B, the TF motif deviations are significantly different in at least three comparisons between acute infection and healthy controls, and at least two comparisons between convalescent and healthy controls (adjust p value ≤ 0.05). The differential gene score analysis was performed using the “getMarkerFeatures” function in ArchR with Wilcoxon Rank Sum test. Genes with FDR ≤ 0.1 were used as input in gene set over-representation analysis for enrichment of Blood Transcriptomic Modules with hypergeometric test. The tracks of antiviral regions were generated using the “plotBrowserTrack” function in ArchR.

### Correlation network

The correlation networks were computed using four data types: 1) transcriptomics, in which the average scores of BTM clusters in each cell type in each sample were used as features; 2) epigenomics, in which the average deviations of TF motif clusters in each cell type in each sample were used as features; 3) proteomics/CyTOF, in which the average level of proteins in each cell type in each sample were used as features; 4) Plasma cytokine levels in monocytes and dendric cells; 5) IFNa2 levels. The BTM score in each cell was calculated as the geometric mean of the expression of all the measured genes in the BTM gene sets. The BTM scores were then aggregated by each sample and each cell type, and hierarchical clustering (Euclidean distance, ward.D2 agglomeration method) was applied to identify BTM clusters. The TF motif clusters were identified in a similar way. Spearman’s rho correlation was used in all the correlation calculations. The correlation networks were identified and visualized using igraph ^96^ implemented in the ggraph package ^97^. Fruchterman-Reingold layout was used.

### Bulk RNA-seq

Blood was collected into Tempus Blood RNA tubes (Applied Biosystems) and the RNA was extracted using the MagMAX for Stabilized Blood Tubes RNA Isolation Kit, compatible with Tempus Blood RNA Tubes (ThermoFisher Scientific). RNA quality was assessed using a TapeStation 4200 (Agilent) and then 200 nanograms of total RNA was used as input for cDNA synthesis and library preparation using the KAPA RNA HyperPrep kit with RiboErase (HMR) Globin (Roche) according to the manufacturer’s instructions. Libraries were validated by capillary electrophoresis on a TapeStation 4200 (Agilent), pooled at equimolar concentrations, and sequenced with PE100 reads on an Illumina NovaSeq 6000, yielding ∼60 million reads per sample on average.

### Bulk RNA-seq analysis

Alignment was performed using STAR version 2.7.3a ^98^ and transcripts were annotated using a composite genome reference which included GRCh38 Ensembl release 100 and SARS-CoV-2 (GCF_009858895.2,ASM985889v3). Transcript abundance estimates were calculated internal to the STAR aligner using the algorithm of htseq-count ^99^. ENSEMBL IDs were filtered to remove low/non-expressed transcripts (<5 reads in >50% of samples). Gene-level counts were created by averaging counts from all ENSEMBL IDs mapping to the same gene symbol (IDs mapping to multiple symbols were discarded), using the bioMart package. Gene counts were normalized using the estimateSizeFactors function of DESeq2. To evaluate monocyte signatures expression during infection, scores for each signature were computed as the average of all genes within the signature. All analysis was performed in R 4.1.1. if not stated differently. Figures were generated using ggplot2 ^100^, and Complexheatmap ^101^.

## List of Supplementary materials

### Supplementary Figure legends

**Supp Figure 1.**
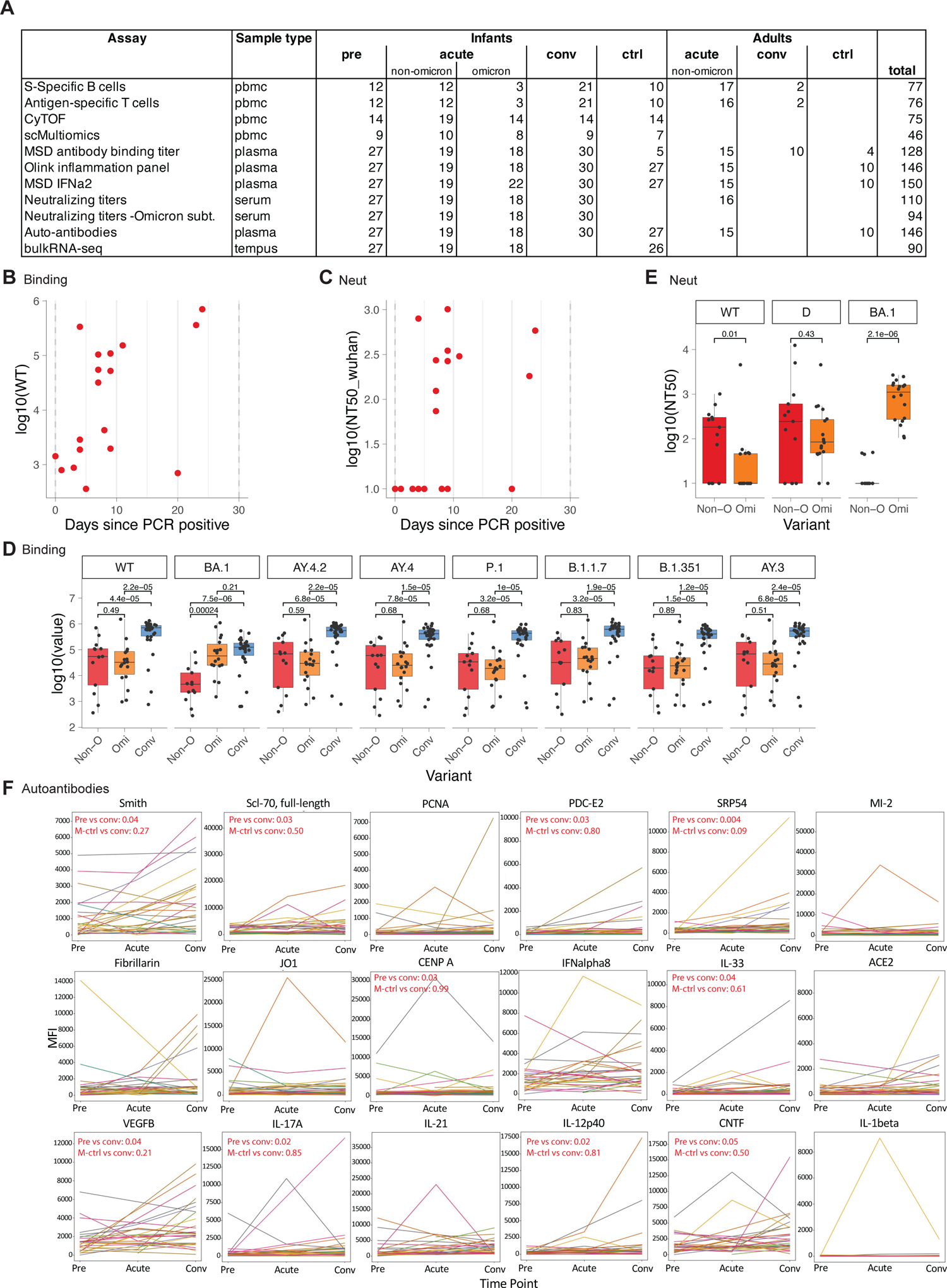
Humoral response to COVID-19 infection in infants, related to **Figure 1** A) Summary of the conducted assays. B,C) Kinetics of developing binding (B) and neutralizing (C) antibody response in infants with COVID-19. D,E) Comparison of binding (D, Non-Omi n = 13, Omi n = 18, Conv n = 30) and neutralizing (E, Non-O n = 13, Omi n = 18) titers between different variants in infants. E) Kinetics of specific autoantibodies which increased during or after COVID-19. Initial statistical comparisons were conducted in a paired fashion with the Wilcoxon signed rank test (Pre vs. Conv, n = 27) and Benjamini-Hochberg correction. Validation tests (M-ctrl vs. Conv) were conducted using Wilcoxon rank sum test (conv n = 30, m-ctrl n = 27).

**Supp Figure 2.**
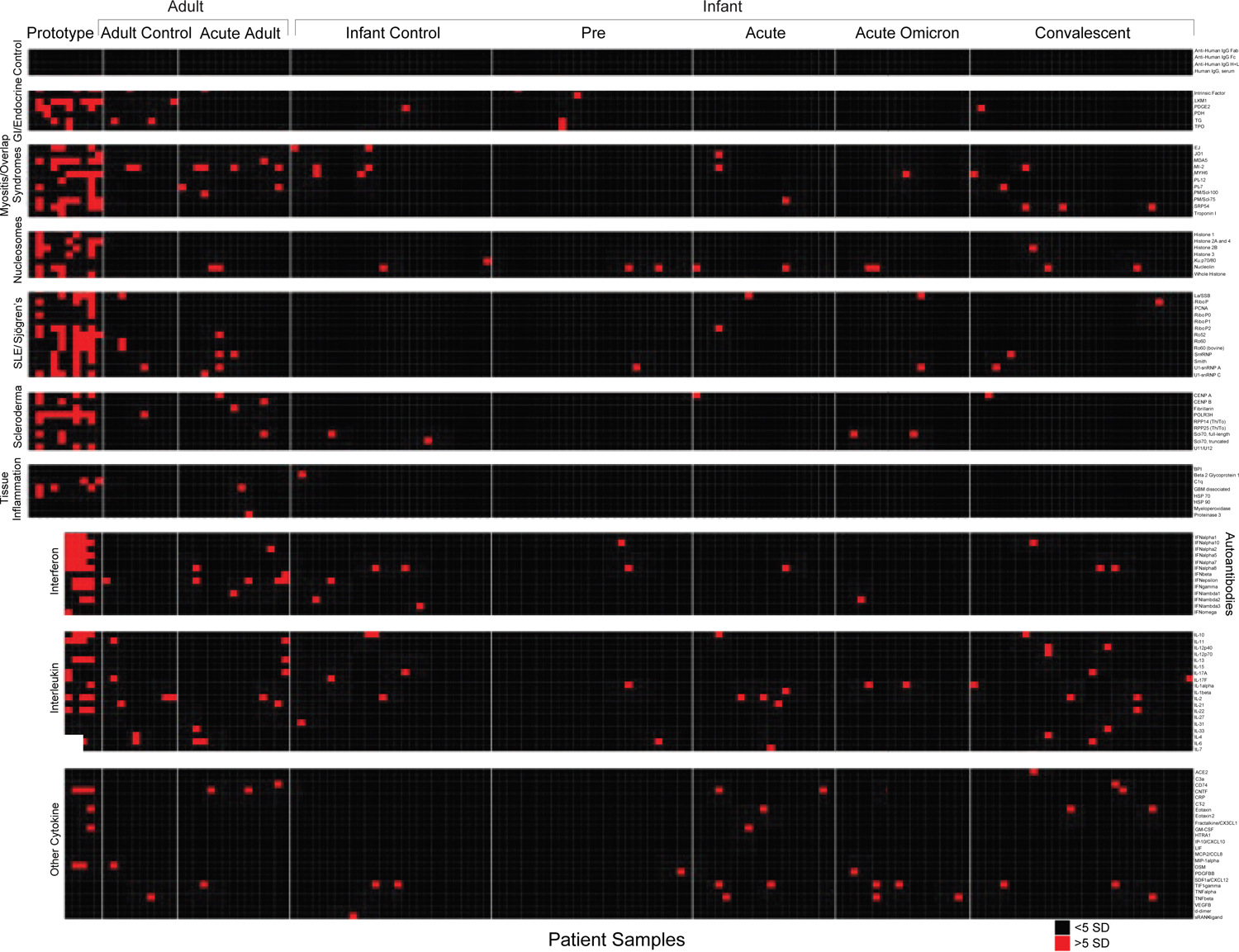
Prevalence of autoantibodies in infants with COVID-19, related to **Figure 1** Heatmap depicting plasma IgG antibodies against the indicated autoantigens and cytokines and chemokines. Prototypes (n=15), adult controls (n=10), adults with acute COVID-19 infection (n=15), infant controls who have not had COVID-19 (n=27), and longitudinal samples from infants who had COVID-19 (pre-infection (n=27), acute infection (n=19), acute omicron infection (n=18), and convalescent samples (n=30)) are shown. Prototypes are positive control samples from patients with known autoimmune disorders. Colors indicate autoantibodies with MFI >5 SD (red) or <5 SD (black) above average for healthy infants. MFIs <3000 were excluded.

**Supp Figure 3.**
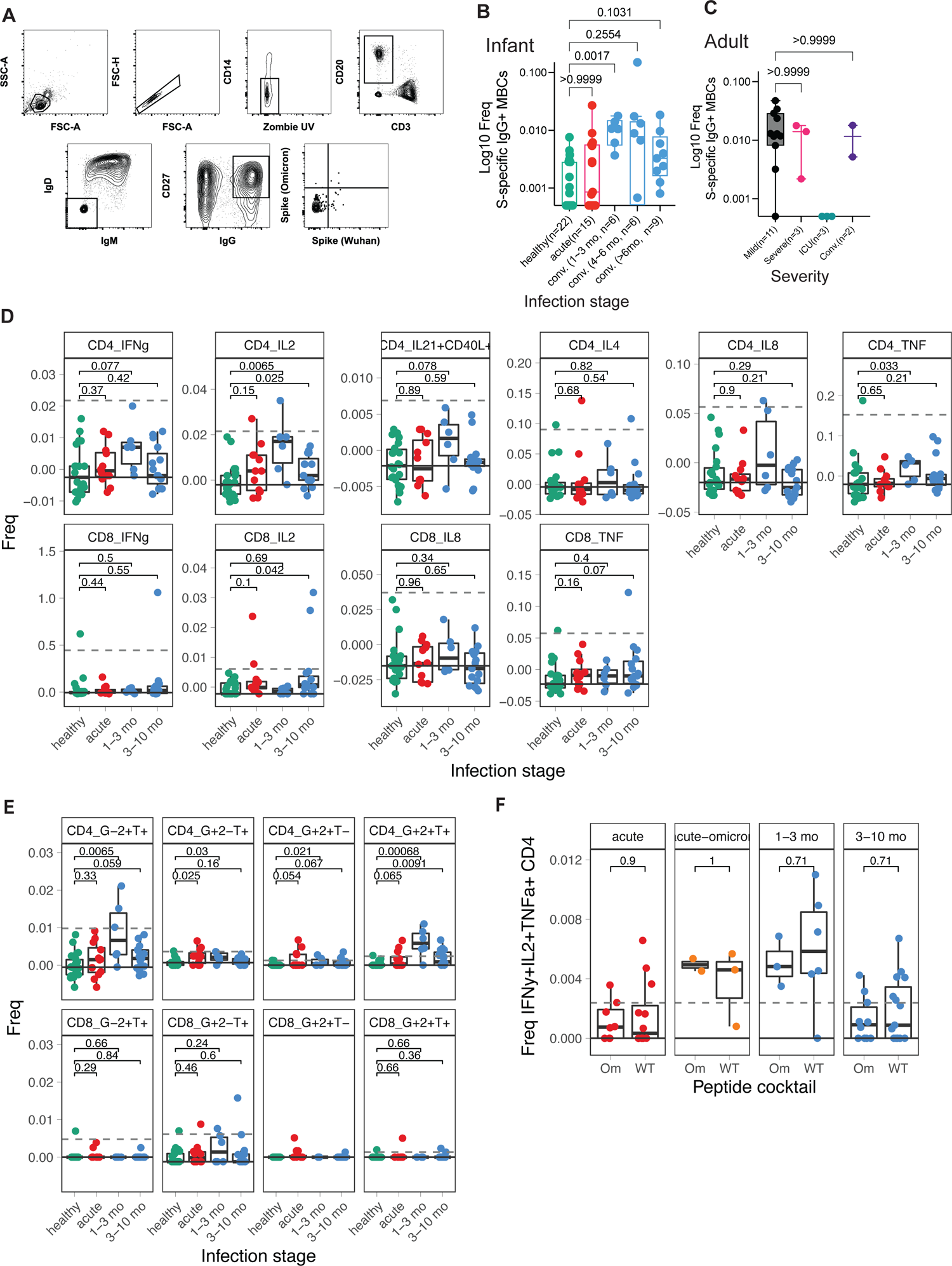
Memory B and T cell response, related to**Figure 2** A) Gating strategy used for SARS-CoV-2 spike specific IgG+ memory B cell staining and single-cell sorting. Gating was on singlets that were CD20+ and CD3-CD14-IgM-IgD-CD27low/+ IgG+. Sorted cells were Wuhan spike-AlexaFluor 488+ and/or Omicron spike-BV421+. B) The percentage of SARS-CoV-2 spike-specific IgG+ memory B cells in healthy, acute, and convalescent infant individuals. The sample number for each group is indicated in brackets. C) As in (B), the percentage of SARS-CoV-2 spike-specific IgG+ memory B cells in adult individuals with mild, severe, and ICU symptoms and in adult convalescent individuals. The sample number for each group is indicated in brackets. D-F) T cells were stimulated with overlapping peptides against WT (D-F) and Omicron (F) variants. Cytokine production was determined via flow cytometry. D) Box plot showing the fraction of responding T cells at different infection stages. E) Box plot showing the fraction of multifunctional T cells at different infection stages. F) Comparison of the multifunctional T cell response after stimulation with WT and Omicron (Om) peptides. Statistical comparisons were conducted with Wilcoxon rank sum test. Solid line indicates median healthy response; dashed line indicates 3x median healthy response.

**Supp Figure 4.**
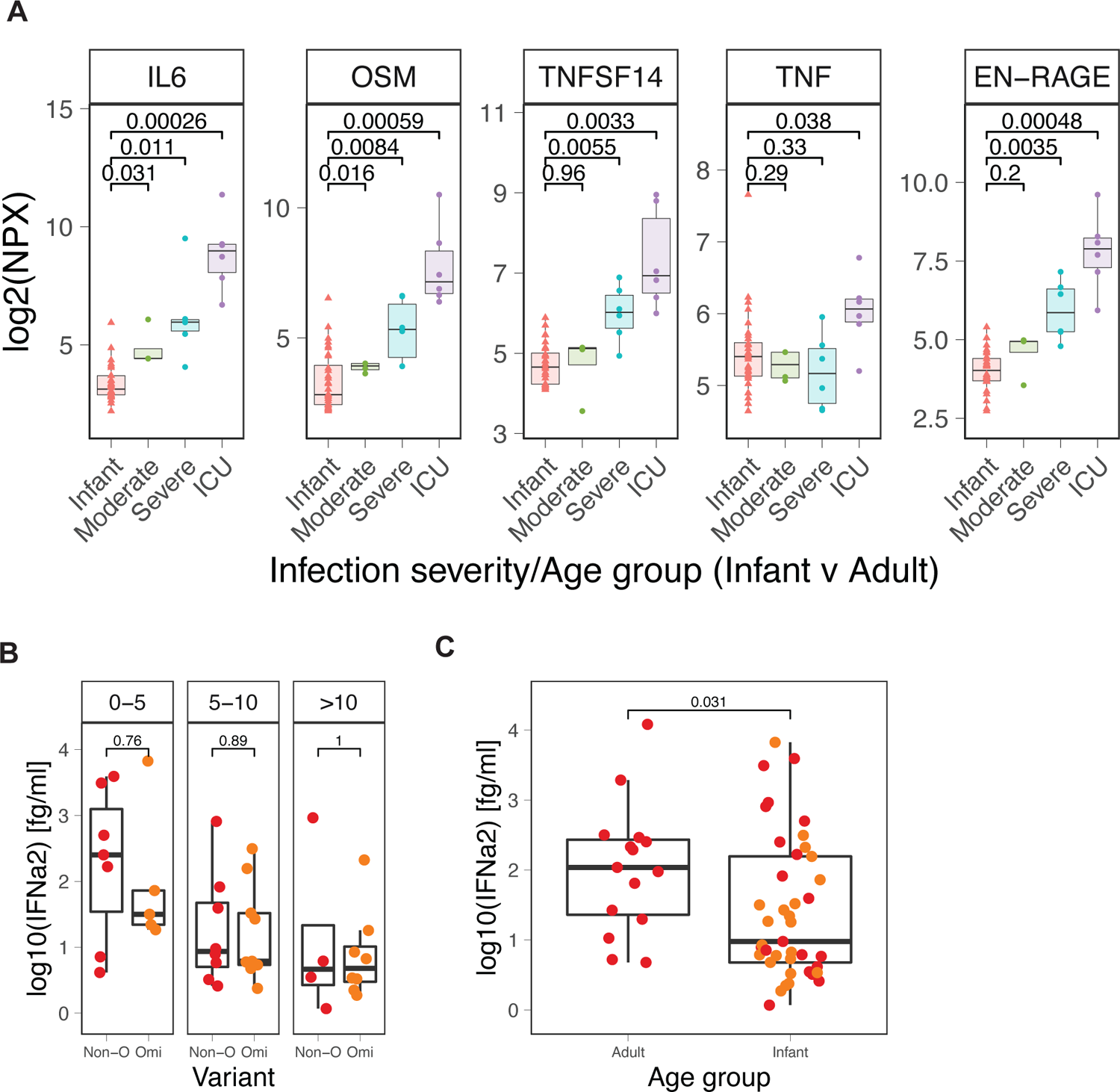
Cytokine response to COVID-19 infection in infants, related to **Figure 3** A) Comparison of key inflammatory mediators during COVID-19 infection in infants and adults, stratified by severity. Infant infection was overall mild or asymptomatic. B,C) Comparison of plasma IFN⍺2 levels between Non-Omicron (Non-O, n = 19) and Omicron (Omi, n = 22) infected infants (B), and infants (n = 41) and adults (n = 15) (C). Statistical comparisons were conducted with the unpaired t-test (A) and Wilcoxon rank sum test (B,C).

**Supp Figure 5.**
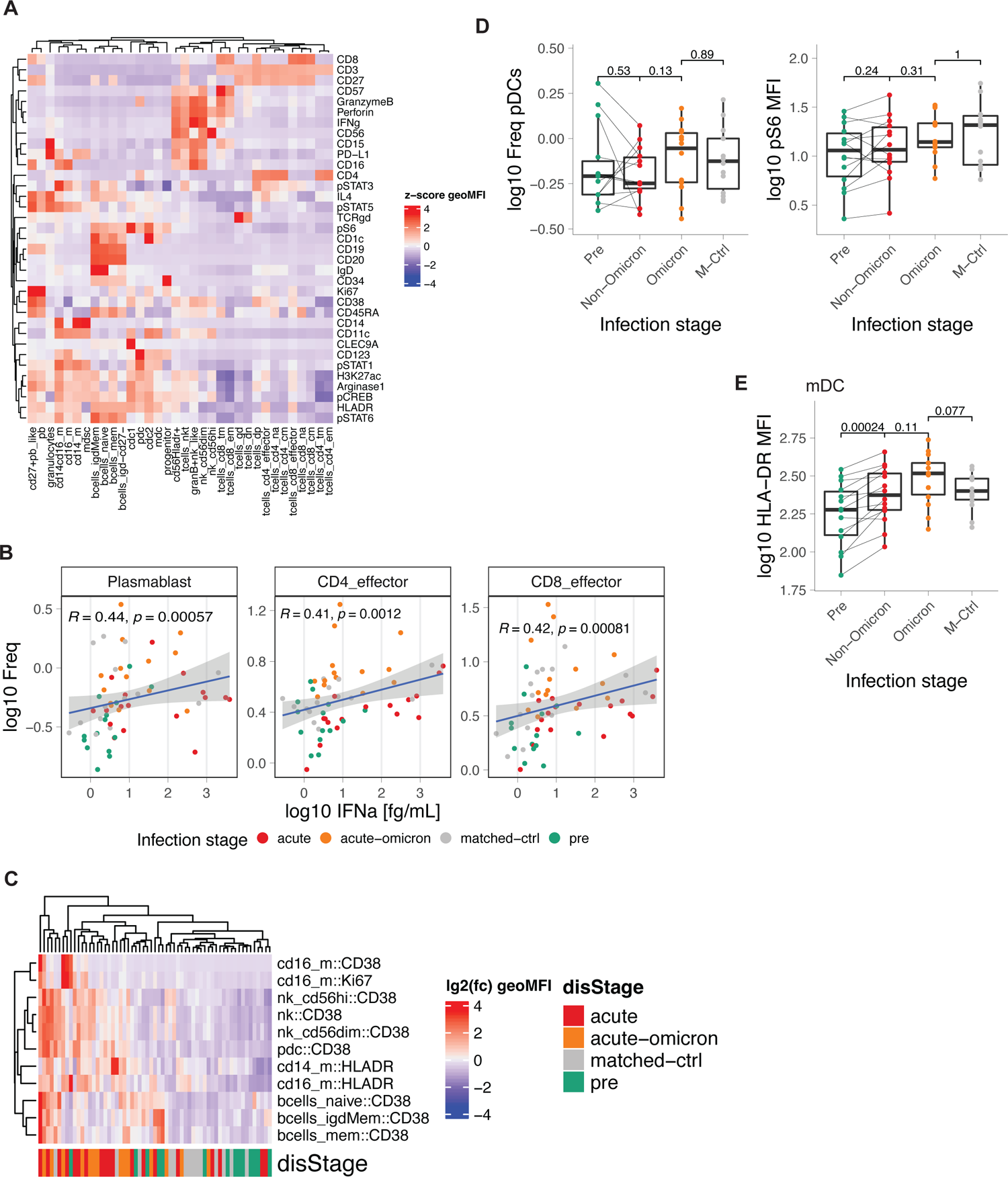
Cellular immune response to COVID-19 infection, related to **Figure 4** A) Heatmap showing the expression of CyTOF markers in all manually gated subsets. B) Correlation analysis between the frequency of indicated cell types (y-axis) and plasma IFN⍺2 levels (x-axis). C) Heatmap showing the expression levels of significantly changed markers in healthy and infected samples. Colors indicate the infection stage. D) Boxplots showing the frequency of pDCs as a proportion of total CD45+ cells (left) and the expression of pS6 in pDCs for different infection stages. E) Boxplots showing HLA-DR expression on mDCs for different infection stages. Correlation analyses were conducted using Spearman correlation.

**Supp Figure 6.**
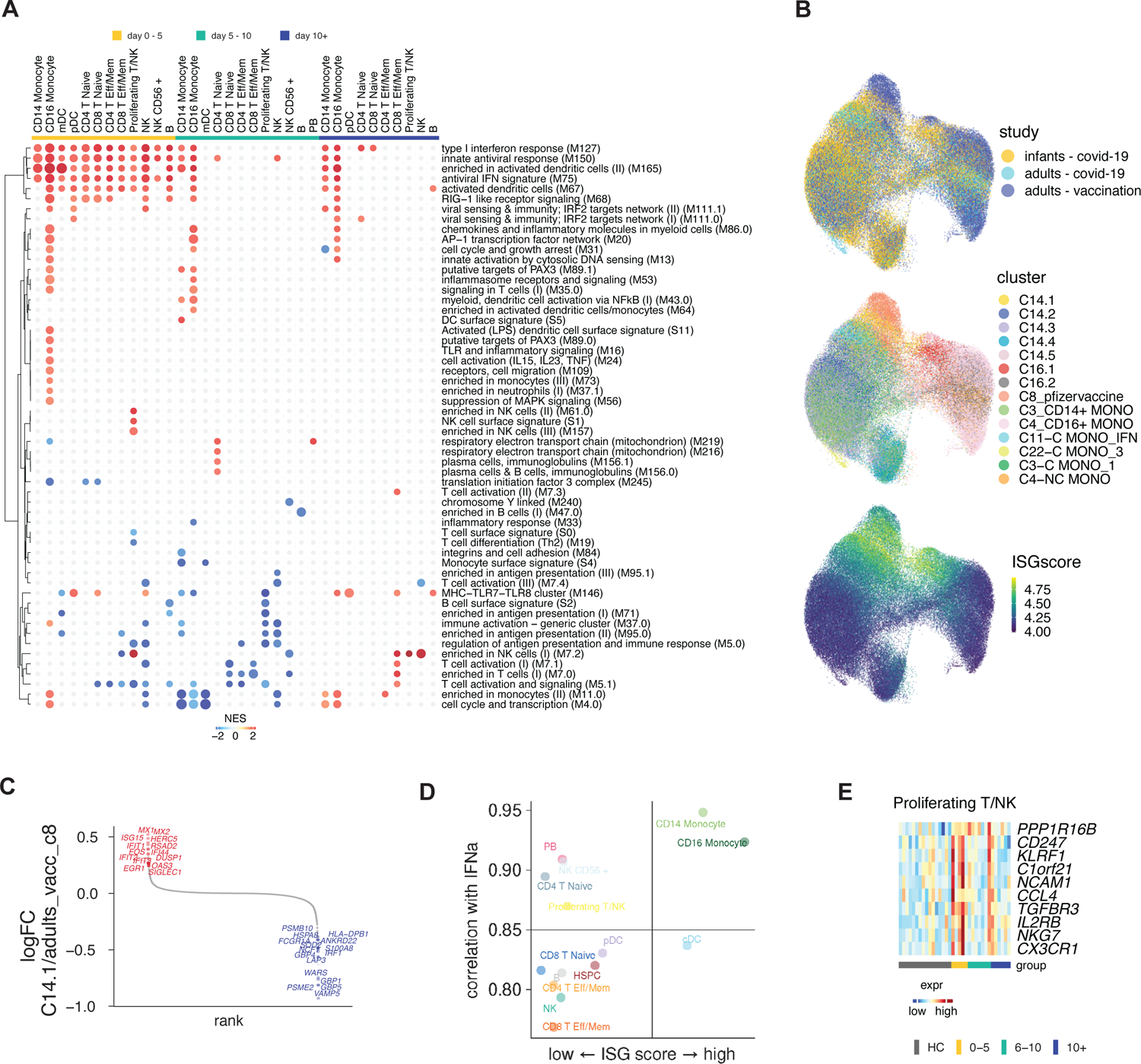
Single-cell multi-omics analysis of immunity to COVID-19 infection in infants, related to **Figure 5** A) Full ring plot from Figure 5c. Pairwise comparison of genes from healthy (n = 16) and COVID-19–infected infants at different times during acute infection (D0-5: n = 5, D5-10: n=7, D10+: n=6) was conducted for each cluster. DEGs were analyzed for the enrichment of BTMs. Ring plot shows an abridged representation of enriched pathways in each cluster. Size indicates the number of samples with enrichment; colors indicate the normalized enrichment score. B) UMAP representation of the integrated analysis of monocyte clusters from this study and from adult COVID-19 patients ^21^ and adult subjects immunized with the COVID-19 vaccine ^30^. Colors indicate the study origin of cells (top), the cell cluster (middle), and the expression of ISGs (bottom). C) DEGs determined between infant C14.1 and adult COVID-19-infection C11 monocyte clusters are plotted and ranked by fold change. C) DEGs determined between infant C14.1 and adult vaccination C8 monocyte clusters are plotted and ranked by fold change. D) Correlation analysis between plasma IFN⍺2 levels and average ISG levels in each cell type. E) Heatmap showing expression of NK cell activation genes enriched in NK cells in Figure 5c (green box).

**Supp Figure 7.**
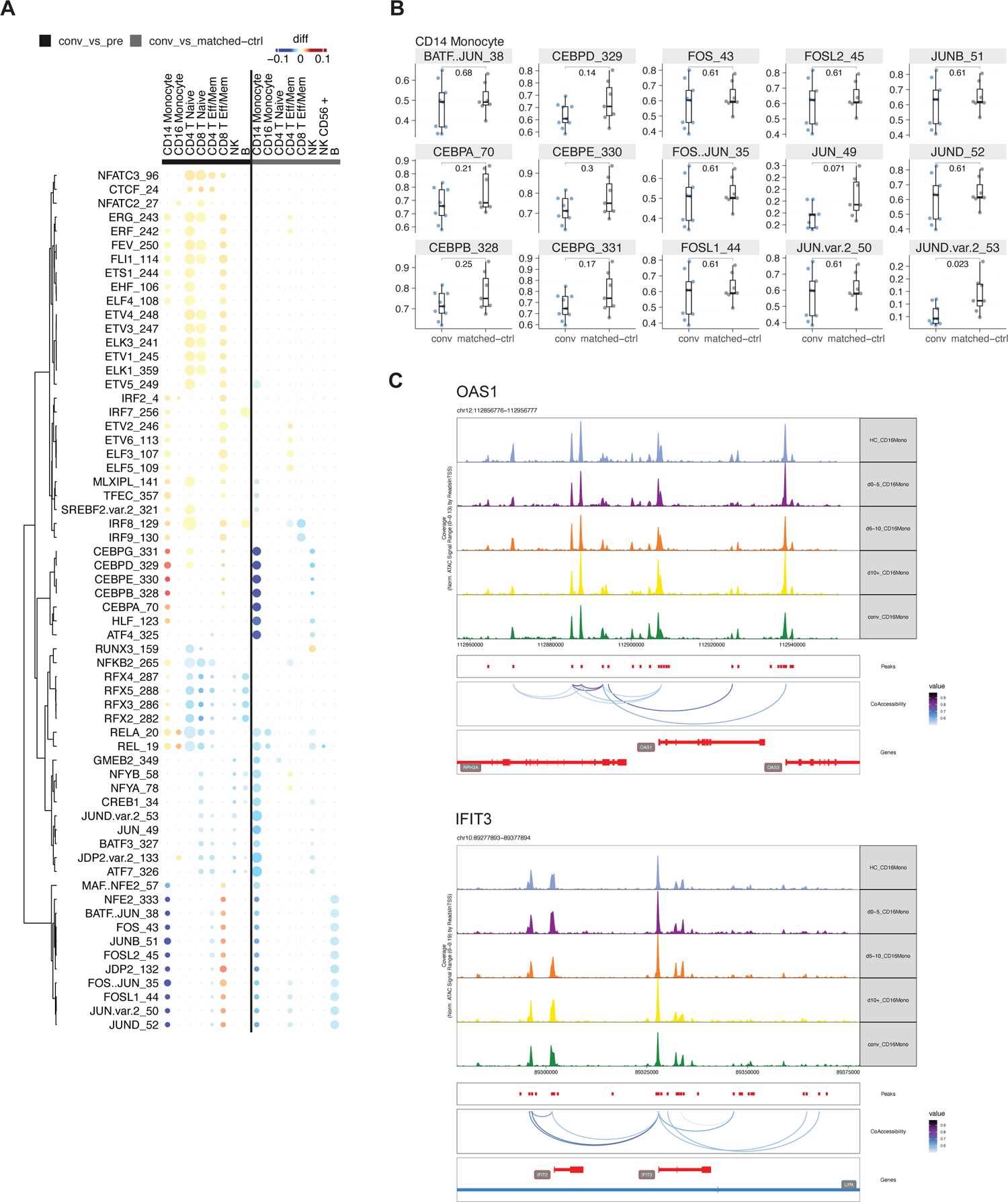
Single-cell epigenomic analysis of immunity to COVID-19 infection in infants, related to **Figure 6** A) Pairwise comparison of TF motif accessibility between convalescent and pre or matched-ctrl samples was conducted for each cluster. Color indicates difference in TF accessibility; non-significant changes (FDR>=0.0001 or changed in less than two subjects) are grey. Size indicates the number of samples with significant change. (pre: n = 9, matched-ctrl: n = 7, Conv: n = 9) B) Box plot showing sample-level accessibility of selected TFs from (A) in CD14+ monocytes. C) Gene tracks showing chromatin accessibility at indicated time points in CD16+ monocytes.

DataS1 - Clinical Information

DataS2 - Autoantibody panel

DataS3 - Flow cytometry and CyTOF information

