## Supplementary material for "Systems biological assessment of the temporal dynamics of immunity to a viral infection in the first weeks and months of life": DataS1 - Clinical tables

**Infant Demographics Table**

| | Controls (n = 27) | Cases (n = 54) |
|---|---|---|
| Female | 11 | 24 |
| Male | 16 | 30 |
| Hispanic | 3 | 3 |
| Not Hispanic | 24 | 51 |
| White | 19 | 42 |
| Black/African American | 4 | 8 |
| White, Black/African American | 4 | 2 |
| White, Other | 0 | 1 |
| American Indian/Native Alaskan | 0 | 1 |
| Pre-Omicron | NA | 32 |
| Omicron | NA | 22 |
| Age in Months (Mean) | 14 (Age at specimen) | 12 (Age at first positive swab) |
| Age in Months (Median) | 12 (Age at specimen) | 9 (Age at first positive swab) |
| Age in Months (Range) | 5-39 (Age at specimen) | 1-47 (Age at first positive swab) |
| Symptomatic Around First Positive Swab (Any Symptom) | NA | 50 |
| Asymptomatic Around First Positive Swab (Any Symptom) | NA | 4 |
| If Symptomatic – Was Cough/Fever Present? | NA | 41 of 50 |

**Adult Demographics Table**

| | Controls (n = 10) | Cases (n = 48) |
|---|---|---|
| Female | 5 | 25 |
| Male | 5 | 23 |
| Hispanic | 1 | 1 |
| Not Hispanic | 7 | 35 |
| No data available | 2 | 12 |
| White | 8 | 14 |
| Black/African American | 0 | 25 |
| Other | 2 | 0 |
| No data available | 0 | 9 |
| Pre-Omicron | NA | 48 |
| Omicron | NA | 0 |
| Age in Years (Mean) | 52 (Age at first positive swab) | 57 (Age at specimen) |
| Age in Years (Median) | 53 (Age at first positive swab) | 59 (Age at specimen) |
| Age in Years (Range) | 25 – 90 (Age at first positive swab) | 24 – 85 (Age at specimen) |
| Mild and moderate disease | NA | 19 |
| Severe disease | NA | 20 |
| ICU | NA | 9 |
