## Supplementary material for "Systems biological assessment of the temporal dynamics of immunity to a viral infection in the first weeks and months of life": DataS2 - Autoantibody antigen list

**DataS3**

**CyTOF antibodies**

| Metal label | Target | Source | Clone | Catalog# | Lot # |
|---|---|---|---|---|---|
| 89Y | CD66b | BioLegend | G10F5 | 305102 | B289360 |
| 102-110Pd | barcodes | Fluidigm | | 201060 | |
| 113In | CD57 | BioLegend | HNK-1 | 359602 | B256178 |
| 140Ce | Beads | Fluidigm | | 201078 | 2108040-25 |
| 141Pr | HLA-DR | BioLegend | L243 | 307651 | B293412 |
| 142Nd | CD19 | Fluidigm | HIB19 | 3142001B | 2110583-20 |
| 143Nd | CD127 | Fluidigm | A019D5 | 3143012B | 2109346-24 |
| 144Nd | IL4 | Fluidigm | MP4-25D2 | 3144010B | 1641910 |
| 145Nd | CD4 | Fluidigm | RPA-T4 | 3145001B | 2109320-22 |
| 146Nd | IgD | Fluidigm | IA6-2 | 3146005B | 2112420-15 |
| 147Sm | CD20 | Fluidigm | 2H7 | 3147001B | 2108087-31 |
| 148Nd | CD34 | Fluidigm | 581 | 3148001B | 3361909 |
| 149Sm | STAT6 | Fluidigm | 18/P-Stat6 | 3149004A | 2631806 |
| 150Nd | pSTAT5 | Fluidigm | 47 | 3150005A | 2103186-01 |
| 151Eu | CD123 | Fluidigm | 6H6 | 3151001B | 2112140-01 |
| 152Sm | CLEC9A | BioLegend | 8F9 | 353802 | B285751 |
| 153Eu | pSTAT1 | Fluidigm | 4a | 3153005A | 0572002 |
| 154Sm | H3K27ac | ActiveMotif | MABI0309 | MABI0309 | 17014 |
| 155Gd | CD27 | Fluidigm | L128 | 3155001B | 3331901 |
| 156Gd | CD45 | Fluidigm | HI30 | 3156010B | 0652006 |
| 157Gd | CD25 | BioLegend | M-A251 | 356102 | B332599 |
| 158Gd | pSTAT3 | HIMC | 4/P-Stat3 | 3158005A | 0132015 |
| 159Tb | CD11c | Fluidigm | Bu15 | 3159001B | 0622021 |
| 160Gd | CD14 | Fluidigm | M5E2 | 3160001B | 2102949-16 |
| 161Dy | Ki-67 | Fluidigm | B56 | 3161007B | 1262010 |
| 162Dy | CD1c | BioLegend | L161 | 331502 | B341969 |
| 163Dy | TCRg/d | BioLegend | B1 | 331202 | B336422 |
| 164Dy | Arginase-1 | Fluidigm | 658922 | 3164012B | 2191531 |
| 165Ho | pCREB | Fluidigm | 87G3 | 3165009A | 1812007 |
| 166Er | CD16 | BioLegend | B73.1 | 360702 | B243320 |
| 167Er | CD38 | Fluidigm | HIT2 | 3167001B | 0902009 |
| 168Er | CD8 | Fluidigm | SK1 | 3168002B | 2103185-01 |
| 169Tm | CD45RA | Fluidigm | HI100 | 3169008B | 2109314-21 |
| 170Er | CD3 | Fluidigm | UCHT1 | 3170001B | 1691904 |
| 171Yb | Granzyme B | Fluidigm | GB11 | 3171002B | 2111006-23 |
| 172Yb | CD15 | Fluidigm | W6D3 | 3172021B | 2103359-16 |
| 173Yb | Perforin | abcam | B-D48 | ab47225 | GR3362878-9 |
| 174Yb | IFNg | BioLegend | 4S.B3 | 502502 | B259482 |
| 175Lu | pS6 | Fluidigm | N7-548 | 3175009A | 3451912 |
| 176Yb | CD56 | Fluidigm | NCAM16.2 | 3176008B | 2151801 |
| 191Ir | DNA1 | Fluidigm | | 201192A | P19K2303 |
| 193Ir | DNA2 | Fluidigm | | 201192A | P19K2303 |
| 209Bi | PD-L1 | Fluidigm | MIH1 | 3209014B | 2102823-01 |

**Antibodies for T cell ICS assay**

| Fluorochrome | Antigen | Vendor | Catalog# | Clone | Reaction | Volume per reaction (μl) |
|---|---|---|---|---|---|---|
| FITC | IL-2 | Biolegend | 500304 | MQ1-17H12 | ICS | 2 |
| PerCP-eF710 | CXCR5 | Invitrogen | 46-9185-42 | MU5UBEE | Stimulation | 2.5 |
| PE | IL-4 | BioLegend | 500810 | MP4-25D2 | ICS | 1 |
| PE-CF594 | CD45RA | BD Biosciences | 565419 | 5H9 | Surface | 2 |
| PE-Cy7 | TNF- α | E-Bioscience | 25-7349-82 | Mab11 | ICS | 0.3 |
| BV421 | CD40L | Biolegend | 310824 | 24-31 | ICS | 2 |
| BV506 | TCR-γδ | Biolegend | 331220 | B1.1 | Surface | 2.5 |
| BV605 | CD4 | Biolegend | 317438 | OKT4 | Surface | 1.5 |
| BV650 | CD3 | BD Biosciences | 563916 | SP34-2 | Surface | 2.5 |
| BV711 | CCR7 | Biolegend | 353228 | G043H7 | Surface | 2 |
| BV785 | CD127 | Biolegend | 351330 | A019D5 | Surface | 2.5 |
| APC | IL-21 | BioLegend | 513008 | 3A3-N2 | ICS | 2.5 |
| A700 | IFN-γ | Biolegend | 502520 | 4S.B3 | ICS | 1 |
| APC-Cy7 | CD25 | Biolegend | 302614 | BC96 | Surface | 2 |
| BUV395 | CXCR3 | BD Biosciences | 565223 | 1C6/CXCR3 | Stimulation | 2.5 |
| BUV563 | CD8 | BD Biosciences | 612914 | RPA-T8 | Surface | 2 |
| BUV737 | CCR6 | BD Biosciences | 612780 | 11A9 | Surface | 2 |
| BUV805 | CD69 | BD Biosciences | 748763 | FN50 | Surface | 2 |

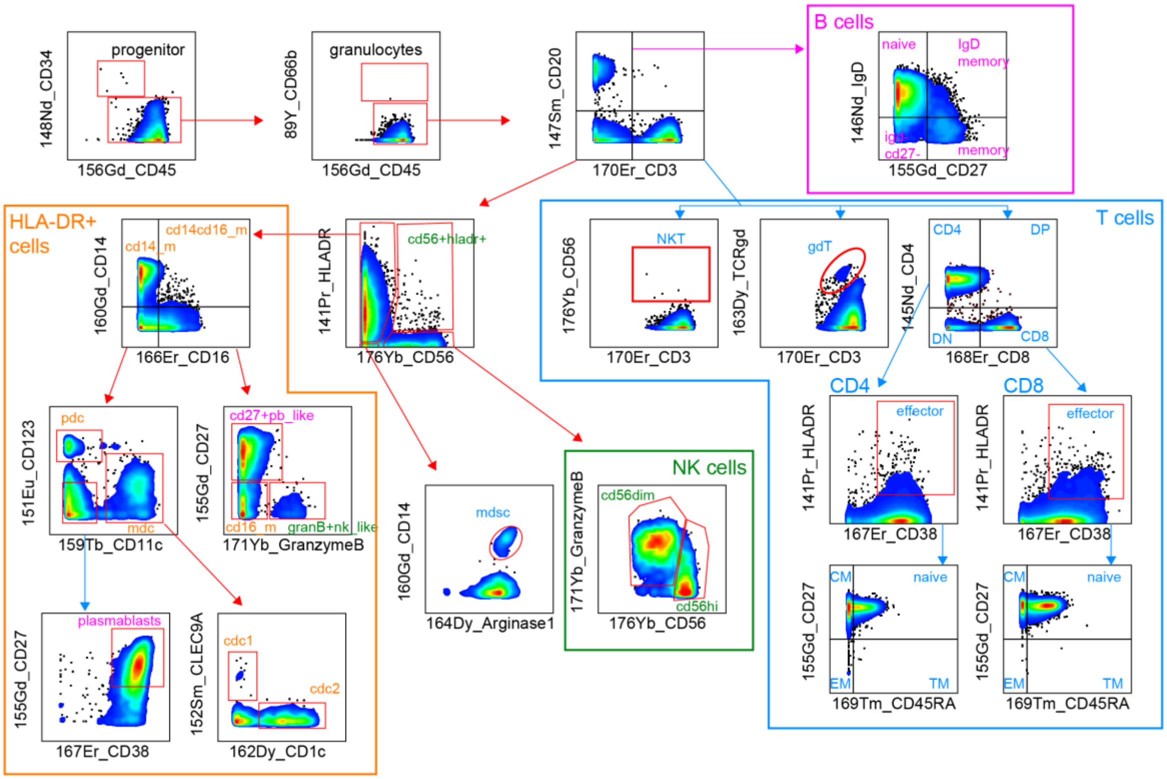


### CyTOF Gating Scheme
