## Supplementary material for "Systems biological assessment of the temporal dynamics of immunity to a viral infection in the first weeks and months of life": DataS3 - Flow and CyTOF antibody tables and gating

**DataS2**

**Antigens auto-antibody panel**

| Antigen | Type | Vendor | Catalogue No |
|---|---|---|---|
| Bare Bead | Control | | |
| Human IgG from serum | Control | Sigma | I4506 |
| Anti-Human IgG Fc specific | Control | Jackson | 109-005-008 |
| Anti-Human IgG H+L | Control | Jackson | 109-005-003 |
| Anti-Human IgG Fab specific | Control | Jackson | 109-005-006 |
| RPP14 (Th/To) | Scleroderma | Origene | TP760291 |
| RPP25 (Th/To) | Scleroderma | Origene | TP303538 |
| CENP A | Scleroderma | Diarect | A16901 |
| CENP B | Scleroderma | Diarect | A12501 |
| Scl-70, full-length | Scleroderma | Diarect | A12401 |
| Scl-70, truncated | Scleroderma | Diarect | A14501 |
| POLR3H | Scleroderma | Origene | TP310633 |
| Fibrillarin | Scleroderma | Prospec | ENZ-566 |
| U11/U12 | Scleroderma | Origene | TP303746 |
| PM/Scl 100 | Myositis/Overlap Syndromes | Diarect | A16001 |
| MDA5 | Myositis/Overlap Syndromes | Diarect | A30001 |
| Troponin I | Myositis/Overlap Syndromes | Prospec | PRO-1269 |
| MYH6 | Myositis/Overlap Syndromes | Origene | TP313673 |
| MI-2 | Myositis/Overlap Syndromes | Diarect | A18101 |
| EJ | Myositis/Overlap Syndromes | Diarect | A11101 |
| JO1 | Myositis/Overlap Syndromes | Diarect | A12901 |
| PL-7 | Myositis/Overlap Syndromes | Diarect | A15601 |
| PL-12 | Myositis/Overlap Syndromes | Diarect | A15701 |
| SRP54 | Myositis/Overlap Syndromes | Diarect | A18401 |
| Ro52 | SLE/Sjogren's | Diarect | A12701 |
| Ro60 (bovine) | SLE/Sjogren's | Diarect | A15501 |
| Ro60 | SLE/Sjogren's | Diarect | A17401 |
| La/SSB | SLE/Sjogren's | Diarect | A12801 |
| Sm/RNP | SLE/Sjogren's | Immunovision | SRC-3000 |
| Smith | SLE/Sjogren's | Immunovision | SMA-3000 |
| U1-snRNP A | SLE/Sjogren's | Diarect | A13101 |

| | | | |
|---|---|---|---|
| U1-snRNP C | SLE/Sjogren's | Diarect | A13201 |
| PCNA | SLE/Sjogren's | Diarect | A15401 |
| Ribo P0 | SLE/Sjogren's | Diarect | A14101 |
| Ribo P2 | SLE/Sjogren's | Diarect | A14301 |
| PDH | GI/Endocrine | Sigma | P7032 |
| PDC-E2 | GI/Endocrine | Diarect | A17901 |
| TPO | GI/Endocrine | Diarect | A12101 |
| TG | GI/Endocrine | Diarect | A12201 |
| Intrinsic Factor | GI/Endocrine | Diarect | A16701 |
| LKM1 | GI/Endocrine | Diarect | A13501 |
| Ku, p70/80 | Nucleosomes | Diarect | A17301 |
| Nucleolin | Nucleosomes | Diarect | A19701 |
| Whole Histone | Nucleosomes | Immunovision | HIS-1000 |
| Histone 1 | Nucleosomes | Immunovision | HIS-1001 |
| Histone 2A and 4 | Nucleosomes | Immunovision | HIS-1002 |
| Histone 2B | Nucleosomes | Immunovision | HIS-1003 |
| Histone 3 | Nucleosomes | Immunovision | HIS-1004 |
| C1q | Tissue Inflammation | Biodesign | A90150H |
| Beta 2 Glycoprotein 1 | Tissue Inflammation | Diarect | A14901 |
| Myeloperoxidase | Tissue Inflammation | Diarect | A18501 |
| Proteinase 3 | Tissue Inflammation | Diarect | A18601 |
| BPI | Tissue Inflammation | Arotec | ATB01-02 |
| GBM dissociated | Tissue Inflammation | Diarect | A16801 |
| HSP 70 | Tissue Inflammation | Stressgen | NSP-555 |
| HSP 90 | Tissue Inflammation | Stressgen | SPP-770 |
| PM/Scl-75 | Myositis/Overlap Syndromes | Diarect | A17001 |
| Ribo P1 | SLE/Sjogren's | Diarect | A14201 |
| IFNalpha1 | Interferon | Prospec | CYT-291 |
| IFNalpha2 | Interferon | R&D | 11101-2 |
| IFNalpha6 | Interferon | MyBioSource | MBS1351081 |
| IFNalpha7 | Interferon | Prospec | CYT-196 |
| IFNalpha8 | Interferon | Sino | 10347-H08H |
| IFNalpha10 | Interferon | Sino | 10349-H08H |
| IFNbeta | Interferon | Peprotech | 300-02BC |
| IFNgamma | Interferon | Peprotech | 300-02 |
| IFNepsilon | Interferon | R&D | 9667-ME-025/CF |
| IFNlambda1 | Interferon | Peprotech | 300-02L |
| IFNlambda2 | Interferon | Peprotech | 300-02K |

| | | | |
|---|---|---|---|
| IFNlambda3 | Interferon | R&D | 5259-IL-025/CF |
| IFNomega | Interferon | R&D | 11395-1 |
| IL-1alpha | Interleukin | Prospec | CYT-253 |
| IL-1beta | Interleukin | Peprotech | 200-01B |
| IL-2 | Interleukin | Peprotech | 200-02 |
| IL-4 | Interleukin | Peprotech | 200-04 |
| IL-6 | Interleukin | Peprotech | 200-06 |
| IL-7 | Interleukin | Peprotech | 200-07 |
| IL-10 | Interleukin | Peprotech | 200-10 |
| IL-11 | Interleukin | Prospec | CYT-214 |
| IL-12p40 | Interleukin | Sino | 10052-H02H |
| IL-12p70 | Interleukin | Peprotech | 200-12 |
| IL-13 | Interleukin | Peprotech | 200-13 |
| IL-15 | Interleukin | Peprotech | 200-15 |
| IL-17A | Interleukin | Peprotech | 200-17 |
| IL-17F | Interleukin | Peprotech | 200-25 |
| IL-21 | Interleukin | Peprotech | 200-21 |
| IL-22 | Interleukin | Peprotech | 200-22 |
| IL-27 | Interleukin | Prospec | CYT-048 |
| IL-31 | Interleukin | Prospec | CYT-625 |
| IL-33 | Interleukin | Peprotech | 200-33 |
| ACE2 | Other Cytokine | Sino | 10108-H05H |
| C3a | Other Cytokine | R&D | 3677-C3-025 |
| CD74 | Other Cytokine | Prospec | PRO-1467 |
| CNTF | Other Cytokine | Prospec | CYT-272 |
| CRP | Other Cytokine | Prospec | PRO-335 |
| CT-2 | Other Cytokine | Prospec | PRO-1578 |
| d-dimer | Other Cytokine | LeeBio | 200-13-0.1 |
| Eotaxin | Other Cytokine | Peprotech | 300-21 |
| Eotaxin 2 | Other Cytokine | Peprotech | 300-33 |
| Fractalkine/CX3CL1 | Other Cytokine | Peprotech | 300-31 |
| GM-CSF | Other Cytokine | Peprotech | 300-03 |
| HTRA1 | Other Cytokine | R&D | 2916-SE-020 |
| IP-10/CXCL10 | Other Cytokine | Peprotech | 300-12 |
| LIF | Other Cytokine | Peprotech | 300-05 |
| MCP-2/CCL8 | Other Cytokine | Peprotech | 300-15 |
| MIP-1alpha | Other Cytokine | Peprotech | 300-08 |
| OSM | Other Cytokine | Peprotech | 300-10 |

| | | | |
|---|---|---|---|
| PDGFBB | Other Cytokine | Peprotech | 100-14B |
| SDF-1a/CXCL12 | Other Cytokine | Peprotech | 300-28A |
| sRANK-ligand | Other Cytokine | Peprotech | 310-01C |
| TIF1-gamma | Other Cytokine | Diarect | A11001 |
| TNFalpha | Other Cytokine | Peprotech | 300-01A |
| TNFbeta | Other Cytokine | Peprotech | 300-01B |
| VEGFB | Other Cytokine | Peprotech | 100-20B |
